# Convalescent plasma for hospitalized patients with COVID-19 and the effect of plasma antibodies: a randomized controlled, open-label trial

**DOI:** 10.1101/2021.06.29.21259427

**Authors:** The CONCOR-1 Study Group, CONCOR-1 writing committee, Philippe Bégin, Jeannie Callum, Erin Jamula, Richard Cook, Nancy M. Heddle, Alan Tinmouth, Michelle P. Zeller, Guillaume Beaudoin-Bussières, Luiz Amorim, Renée Bazin, Kent Cadogan Loftsgard, Richard Carl, Michaël Chassé, Melissa M. Cushing, Nick Daneman, Dana V. Devine, Jeannot Dumaresq, Dean A. Fergusson, Caroline Gabe, Marshall J. Glesby, Na Li, Yang Liu, Allison McGeer, Nancy Robitaille, Bruce S. Sachais, Damon C. Scales, Lisa Schwartz, Nadine Shehata, Alexis F. Turgeon, Heidi Wood, Ryan Zarychanski, Andrés Finzi, Donald M. Arnold, for The CONCOR-1 Study Group

## Abstract

The efficacy of convalescent plasma for COVID-19 is unclear. While most randomized controlled trials have shown negative results, uncontrolled studies have suggested that the antibody content may influence patient outcomes.

We conducted an open-label, randomized controlled trial of convalescent plasma for adults with COVID-19 receiving oxygen within 12 days of respiratory symptom onset. Patients were allocated 2:1 to 500 mL of convalescent plasma or standard of care. The composite primary outcome was intubation or death by 30 days. The effect of convalescent plasma antibodies on the primary outcome was assessed by logistic regression.

The trial was terminated at 78% of planned enrollment after meeting stopping criteria for futility. 940 patients were randomized and 921 patients were included in the intent-to-treat analysis. Intubation or death occurred in 199/614 (32.4%) in the convalescent plasma arm and 86/307 (28.0%) in the standard of care arm; relative risk (RR) 1.16 (95% confidence interval (CI) 0.94-1.43; p=0.18). Patients in the convalescent plasma arm had more serious adverse events (33.4% vs. 26.4%; RR=1.27, 95% CI 1.02-1.57, p=0.034). The antibody content significantly modulated the therapeutic effect of convalescent plasma. In multivariate analysis, each standard log increase in neutralization or antibody-dependent cellular cytotoxicity independently reduced the potential harmful effect of plasma (OR=0.74; 0.57-0.95 and OR=0.66; 0.50-0.87, respectively), while IgG against the full transmembrane Spike protein increased it (OR=1.53, 95% CI 1.14-2.05).

Convalescent plasma did not reduce the risk of intubation or death at 30 days among hospitalized patients with COVID-19. Transfusion of convalescent plasma with unfavourable antibody profiles may be associated with worse clinical outcomes compared to standard care.

**Trial registration:** CONvalescent Plasma for Hospitalized Adults With COVID-19 Respiratory Illness (CONCOR-1); NCT04348656; https://www.clinicaltrials.gov/ct2/show/NCT04348656

## Introduction

The immune response after SARS-CoV-2 infection results in the formation of antibodies that can interfere with viral replication and infection of host cells in over 95% of patients.^1^ Based on prior experience in other viral infections,^2^ the use of convalescent plasma has been proposed as a therapeutic form of passive immunization for patients with acute Coronavirus Disease 2019 (COVID-19).^3, 4^ Early in the pandemic, several small randomized trials found no difference in clinical outcomes.^5–8^ In the United States, an Extended Access Program outside of a controlled trial led to the use of convalescent plasma in over half a million patients. Data from these patients showed that the transfusion of plasma with high anti-SARS-CoV-2 antibody level was associated with a lower risk of death in non-intubated patients compared with lower antibody levels; however, this study lacked a control group.^9^ The RECOVERY trial was a large randomized trial in 11,558 hospitalized patients, which found the risk of death following administration of high-titer plasma was not different from standard of care.^10^

The Convalescent Plasma for COVID-19 Respiratory Illness (CONCOR-1) trial was a multi-center, international, open-label, randomized controlled trial designed to assess the effectiveness and safety of COVID-19 convalescent plasma in hospitalized patients. The trial used plasma collected from four blood suppliers with a range of anti-SARS-CoV-2 antibody levels. The variability in antibody titers allowed for a characterization of the effect-modifying role of functional and quantitative antibodies on the primary outcome (intubation or death at 30 days).

## Methods

### Trial Design and Oversight

CONCOR-1 was an investigator-initiated, multi-center, open-label, randomized controlled trial conducted at 72 hospital sites in Canada, the United States, and Brazil.^11^ Eligible patients were randomly assigned to receive either convalescent plasma or standard of care. The study was approved by Clinical Trials Ontario (Research Ethics Board of Record: Sunnybrook Health Sciences Centre), project #2159; the Quebec Ministry of Health and Social Services multicenter ethics review (REB of Record: Comité d’éthique de la recherche du CHU Sainte-Justine), project #MP-21-2020-2863; the Weil Cornell Medicine General Institutional Review Board, protocol number 20-04021981; as well as the Comissão Nacional de Ética em Pesquisa, approval 4.305.792. Regulatory authorization was obtained from Health Canada (Control # 238201) and the United States Food and Drug Administration (IND 22075). The trial was registered at clinicaltrials.gov (NCT04348656). An Independent Data Safety Monitoring Committee performed trial oversight and made recommendations following review of safety reports planned at every 100 patients and at the planned interim analysis based on the first 600 patients. External monitoring was performed at all sites to assess protocol adherence, reporting of adverse events, and accuracy of data entry. The full details of the study design, conduct, oversight, and analyses are provided in the protocol and statistical analysis plan, which are available online.

### Participants

Eligible participants were ≥16 in Canada or ≥18 years of age in the United States and Brazil who were admitted to the hospital ward with confirmed COVID-19 and who required supplemental oxygen. The availability of ABO-compatible convalescent plasma from donors who had recovered from COVID-19 infection was an eligibility requirement. Exclusion criteria were: more than 12 days from the onset of respiratory symptoms, imminent or current intubation, a contraindication to plasma transfusion, or a plan for no active treatment. Consent was obtained from all donors and participants (or their legally authorized representative).

### Randomization and Intervention

Patients were randomized in a 2:1 ratio to receive convalescent plasma or standard of care using a secure, concealed, computer-generated, web-accessed randomization sequence (REDCap). Randomization was stratified by site and age (<60 and ≥ 60 years) with allocation made with permuted blocks of size 3 or 6. Patients randomized to convalescent plasma received one or two units of apheresis plasma amounting to approximately 500 mL from one or two donors. The plasma was stored frozen and thawed as per standard blood bank procedures and infused within 24 hours of randomization. Patients were monitored by clinical staff for transfusion-related adverse events as per local procedures. Individuals assigned to standard of care received usual medical care as per routine practices at each site. The investigational product was prepared by Canadian Blood Services and Héma-Québec (Canada), New York Blood Center (USA),^12^ and Hemorio (Brazil). Each supplier had different criteria for qualifying convalescent plasma units that were based on the presence of either viral neutralizing antibodies at a titer of >1:160 or antibodies against the receptor binding domain (RBD) of the SARS-CoV-2 Spike protein at a titer of >1:100. In addition, a sample from each plasma donation was tested at reference laboratories after the transfusion for: (1) anti-RBD antibodies (IgM, IgA and IgG) by enzyme-linked immunosorbent assay (ELISA);^13, 14^ (2) viral neutralization by the plaque-reduction neutralization test using live virus;^15, 16^ (3) IgG antibodies binding to the full-length trimeric transmembrane SARS-CoV-2 Spike protein expressed on 293T cells by flow cytometry;^17^ and, (4) Fc-mediated function by an antibody-dependent cellular cytotoxicity (ADCC) assay against the full Spike protein expressed on CEM.NKr cells (see supplement for complete description).^18, 19^ For each plasma unit, the absolute antibody content was defined as the product of the unit volume and the concentration of the antibody (or functional capacity) in the plasma. These calculations were used to estimate the total antibody content from the transfusion of two units.

### Trial Outcomes

The primary outcome was the composite of intubation or death by day 30. Secondary outcomes were: time to intubation or death; ventilator-free days by day 30; in-hospital death by day 90; time to in-hospital death; death by day 30; length of stay in critical care and hospital; need for extracorporeal membrane oxygenation; need for renal replacement therapy; convalescent plasma-associated adverse events; occurrence of ≥3 grade adverse events by day 30 (classification of adverse events was performed using MedDRA (https://www.meddra.org/) and graded by the Common Terminology Criteria for Adverse Events, version 4.03). All transfusion-related adverse events were classified and graded by the International Society for Blood Transfusion definitions (www.isbtweb.org). All patients were followed to day 30, including a 30-day telephone visit for patients who were discharged from hospital. Patients who were in hospital beyond day 30 were followed until discharge for the purpose of determining in-hospital mortality up to day 90.

### Statistical Analysis

The primary analysis was based on the intention-to-treat population, which included all individuals who were randomized and for whom primary outcome data were available. The per-protocol population was comprised of eligible individuals who were treated according to the randomized allocation of the intervention and received two units (or equivalent) of convalescent plasma within 24 hours of randomization.

The effect of convalescent plasma on the composite primary outcome of intubation or death by day 30 was assessed by testing the null hypothesis that the composite event rate was the same under convalescent plasma and standard of care. The relative risk for the primary outcome (convalescent plasma versus standard of care) was computed with a 95% confidence interval. Secondary outcomes were analyzed as described in the statistical analysis plan (see supplementary material). No multiplicity adjustments were implemented for the secondary analyses. Procedures planned for addressing missing data and subgroup analyses are described in the statistical analysis plan (see supplementary appendix). Forest plots were used to display point estimates and confidence intervals across subgroups with interaction tests used to assess effect modification.

The effect-modifying role of antibody content on the primary outcome was assessed via logistic regression controlling for the blood supplier, treatment, and the antibody marker. Antibody markers were log-transformed, centered and then divided by the corresponding standard deviation before being entered into logistic regression models (see statistical analysis plan, supplementary appendix). A multivariate logistic regression model was then fitted adjusting for all four markers. Generalized additive models were used to examine the joint effect of each pair of serologic markers on the primary outcome.^20^

The results from CONCOR-1 were subsequently included in a meta-analysis based on the May 20^th^ 2021 update of the Cochrane systematic review^8^ and known randomised trials published since comparing convalescent plasma to placebo or standard care in patients with COVID-19. These were divided based on whether they used plasma with high antibody titre or not. For each trial, we compared the observed number of deaths at 30 days (or closest available time point prior to a crossover, if applicable) among patients allocated to convalescent plasma or the control group. Summary estimates for relative risk with 95% confidence interval were calculated using random-effects meta-analysis to account for variation in effect size amongst studies. Heterogeneity was quantified using inconsistency index (I^2^) and p-values from the chi-square test for homogeneity.

With a 2:1 randomization ratio, 1200 patients (800 in the convalescent plasma group, and 400 in the standard of care group) were needed to provide 80% power to detect a relative risk reduction of 25% with convalescent plasma for the primary outcome with a 30% event rate under standard of care, based on a two-sided test at the 5% significance level. An interim analysis by a biostatistician unblinded to the allocation of the intervention was planned for when the primary outcome was available for 50% of the target sample. An O’Brien-Fleming stopping rule was employed^21^ to control the overall type-I error rate at 5%. Conditional power was used to guide futility decisions with the nominal threshold of 20% to justify early stopping.

## Results

### Patients

The trial was stopped at the planned interim analysis because the conditional power estimate was 1.6% (below the stopping criterion of 20%). Between May 14^th^ 2020 and January 29^th^ 2021, 940 patients were randomized (Figure 1, eTable 1). Seventeen patients were lost to follow up between discharge and day 30 and two withdrew consent. Baseline demographics were balanced between groups for all study populations (Tables 1, eTable 2, 3). Median age was 69 years, with 59% male and the median time from the onset of any COVID-19 symptom was 8 days (interquartile range, 5 to 10). The majority of participants (84.0%) were receiving systemic corticosteroids at the time of enrolment.

**Figure 1:**
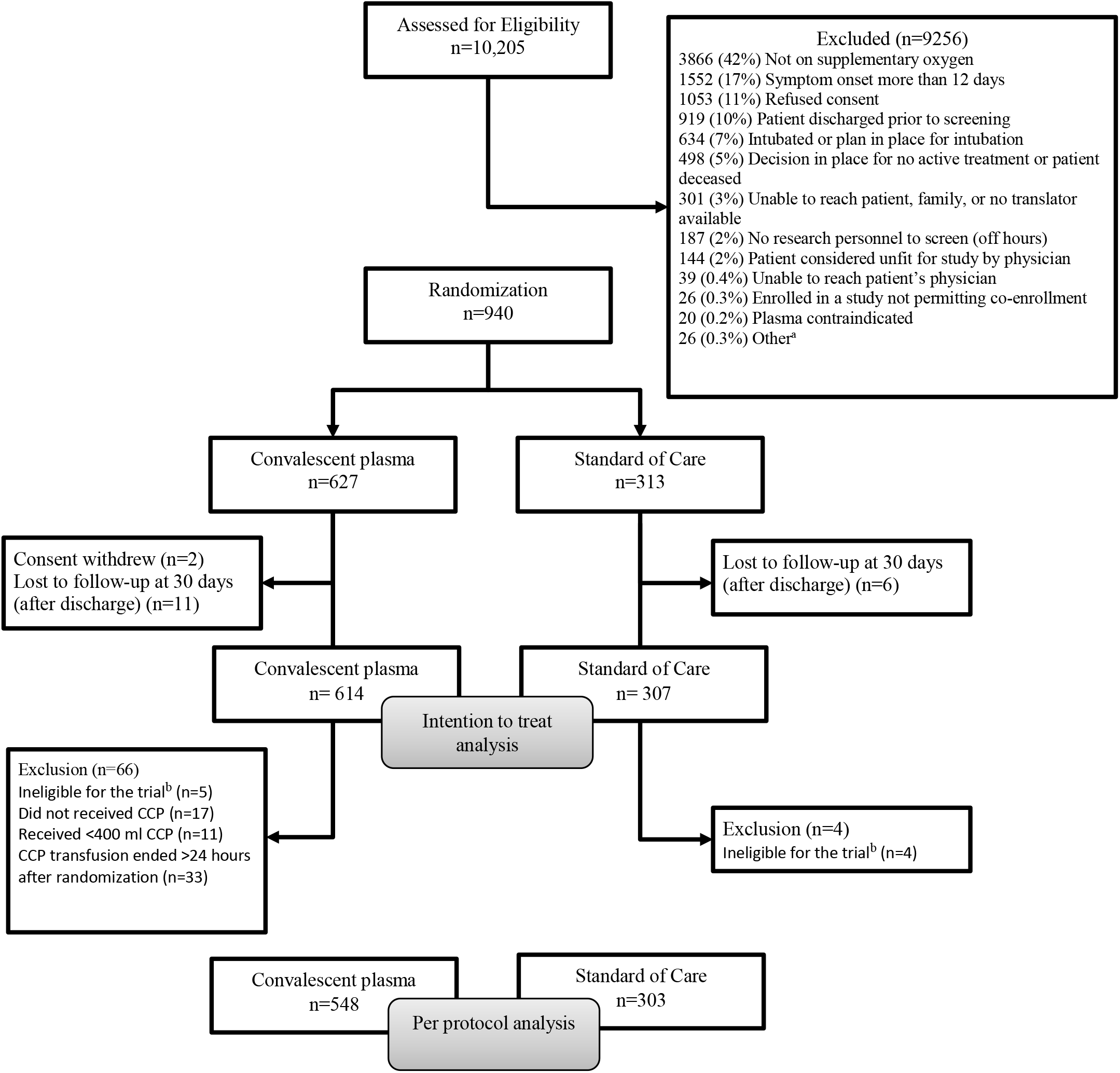
Enrolment, randomization, and follow-up. Patient flow in the CONCOR-1 Study detailing the intention-to-treat population, per-protocol analysis population, and excluded patients. ^a^ Other n=26: <16 years of age (n=13), <18 years of age (n=5), ABO compatible plasma unavailable (n=5), other (n=3) ^b^ Includes not receiving supplemental oxygen at the time of randomization (but on oxygen at screening), and any symptoms onset >12 days prior to randomization for protocol version 5.0 or earlier. CCP: COVID-19 Convalescent Plasma.

**Table 1:** Characteristics of the study population at baseline (excluding two patients who withdrew consent). Categorical data presented as number (percentage) and continuous variables as mean ± standard deviation and median (interquartile range).

| Characteristic | Convalescent Plasma<br>n=625 | Standard of Care<br>n=313 | Overall<br>n=938 |
| --- | --- | --- | --- |
| **Age - yr** | 67.7±16.0; 69 (58,80) | 67.1±14.8; 68 (58, 78) | 67.5±15.6; 69 (58, 79) |
| ≥60 years | 438 (70.1) | 218 (69.6) | 656 (69.9) |
| **Sex** |  |  |  |
| Male | 369 (59.0) | 185 (59.1) | 554 (59.1) |
| **Pregnant at randomization** | 4 (0.6) | 1 (0.3) | 5 (0.5) |
| **Ethnicity** |  |  |  |
| White | 305 (48.8) | 153 (48.9) | 458 (48.8) |
| Asian | 104 (16.6) | 46 (14.7) | 150 (16.0) |
| Hispanic or Latino | 34 (5.4) | 9 (2.9) | 43 (4.6) |
| Black | 25 (4.0) | 11 (3.5) | 36 (3.8) |
| Other | 38 (6.1) | 28 (8.9) | 66 (7.0) |
| Unknown | 119 (19.0) | 66 (21.1) | 185 (19.7) |
| **ABO blood group** |  |  |  |
| O | 270 (43.2) | 113 (36.1) | 383 (40.8) |
| A | 235 (37.6) | 121 (38.7) | 356 (38.0) |
| B | 89 (14.2) | 57 (18.2) | 146 (15.6) |
| AB | 31 (5.0) | 22 (7.0) | 53 (5.7) |
| **BMI – kg/m<sup>2</sup>** | 30.0±7.5; 29 (25, 33) | 30.0±7.4; 29 (25, 33) | 30.0±7.4; 29 (25, 33) |
| BMI <30 | 256 (41.0) | 123 (39.3) | 379 (40.4) |
| BMI ≥ 30 | 198 (31.7) | 102 (32.6) | 300 (32.0) |
| Unknown | 171 (27.4) | 88 (28.1) | 259 (27.6) |
| **Presence of comorbidity** |  |  |  |
| Diabetes | 220 (35.2) | 108 (34.5) | 328 (35.0) |
| Cardiac disease | 385 (61.6) | 197 (62.9) | 582 (62.0) |
| Baseline respiratory diseases | 147 (23.5) | 79 (25.2) | 226 (24.1) |
| **Abnormal CT chest or chest x-ray result before randomization** | 563 (90.1) | 266 (85.0) | 829 (88.4) |
| **Medication for other research study at baseline** | 53 (8.5) | 41 (13.1) | 94 (10.0) |
| **Medication for COVID-19 at baseline** |  |  |  |
| Azithromycin | 279 (44.6) | 137 (43.8) | 416 (44.3) |
| Other antibiotics | 405 (64.8) | 186 (59.4) | 591 (63.0) |
| Systemic corticosteroids | 496 (79.4) | 258 (82.4) | 754 (80.4) |
| Antiviral medications | 165 (26.4) | 80 (25.6) | 245 (26.1) |
| Anticoagulants | 355 (56.8) | 180 (57.5) | 535 (57.0) |
| Other COVID-19 Medications | 79 (12.6) | 39 (12.5) | 118 (12.6) |
| **Medication not for COVID-19 at baseline** |  |  |  |
| ACE inhibitor | 85 (13.6) | 63 (20.1) | 148 (15.8) |
| ACE receptor blocker | 77 (12.3) | 47 (15.0) | 124 (13.2) |
| Non-steroidal anti-inflammatory drugs | 77 (12.3) | 52 (16.6) | 129 (13.8) |
| Colchicine | 5 (0.8) | 2 (0.6) | 7 (0.7) |
| Systemic corticosteroids | 61 (9.8) | 35 (11.2) | 96 (10.2) |
| Inhaled corticosteroids | 84 (13.4) | 42 (13.4) | 126 (13.4) |
| Immunomodulatory agents | 22 (3.5) | 18 (5.8) | 40 (4.3) |
| Anticoagulants | 135 (21.6) | 64 (20.4) | 199 (21.2) |
| **Oxygen status at baseline (FiO<sub>2</sub>)** | 49.5±25.2; 40 (30, 65) | 48.8±25.1; 40 (30, 60) | 49.3±25.2; 40 (30, 60) |
| **Time from any symptom onset to randomization (days)** | 8.0±3.8; 8 (5, 10) | 7.8±3.4; 8 (5, 10) | 7.9±3.7; 8 (5, 10) |
| **Time from Covid-19 diagnosis<sup>a</sup> to randomization (days)** | 4.9±3.6; 4 (2, 7) | 5.1±4.4; 4 (2, 7) | 5.0±3.9; 4 (2, 7) |
| **Enrolled in other clinical trials** | 168 (26.9) | 98 (31.3) | 266 (28.4) |

<sup>a</sup>Day of positive Covid-19 test.
BMI: Body mass index; CT: Computed tomography; FiO<sub>2</sub>: Fraction of inhaled oxygen.

### Primary Outcome

In the intention-to-treat population (n=921), intubation or death occurred in 199 (32.4%) of 614 patients in the convalescent plasma group and 86 (28.0%) of 307 in the standard of care group [relative risk (RR) 1.16; 95% confidence interval (CI) 0.94 to 1.43; p=0.18] (Table 2). The time to intubation or death was not significantly different between groups (eFigure 2). In the per-protocol analysis (n=851), intubation or death occurred in 167 (30.5%) of 548 patients in the convalescent plasma group and 85 (28.1%) of 303 patients in the standard of care group (RR=1.09; 95% CI 0.87 to 1.35; p=0.46) (eTable 4).

**Table 2:**
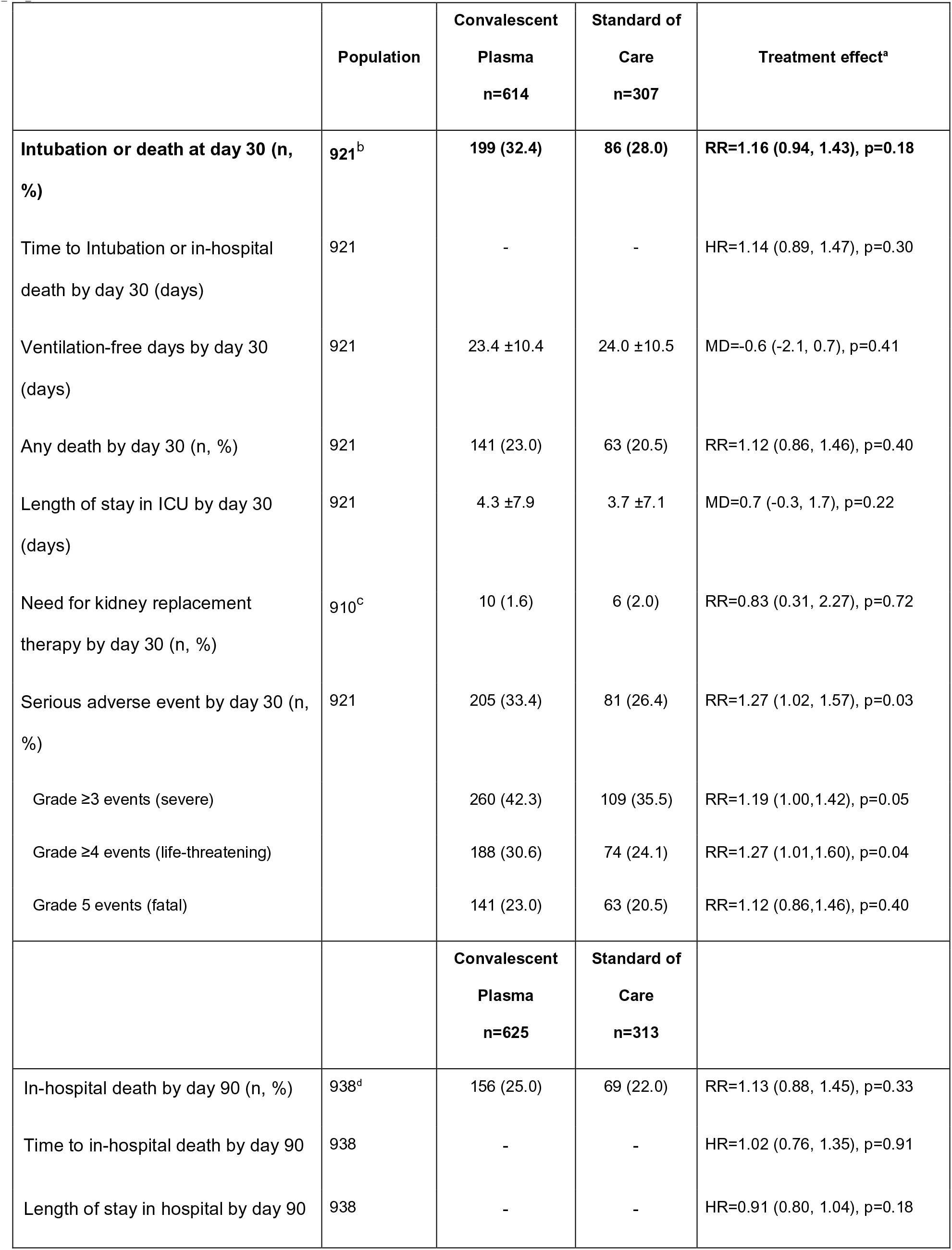
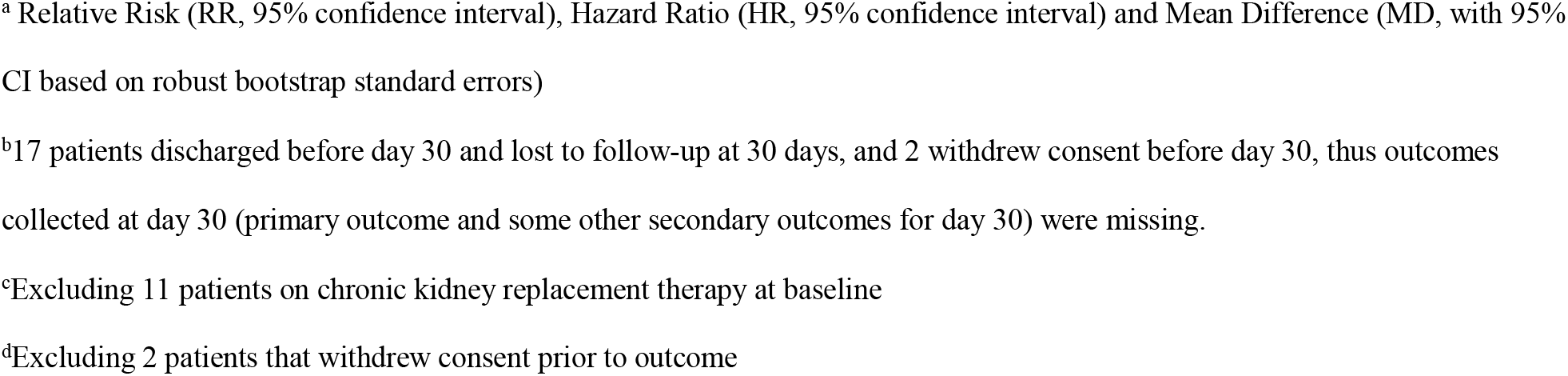
Patient outcomes for the primary and secondary end points for the intention to treat population.

|  | Population | Convalescent<br>Plasma<br>n=614 | Standard of<br>Care<br>n=307 | Treatment effect<sup>a</sup> |
| --- | --- | --- | --- | --- |
| **Intubation or death at day 30 (n, %)** | **921<sup>b</sup>** | **199 (32.4)** | **86 (28.0)** | **RR=1.16 (0.94, 1.43), p=0.18** |
| Time to Intubation or in-hospital death by day 30 (days) | 921 | - | - | HR=1.14 (0.89, 1.47), p=0.30 |
| Ventilation-free days by day 30 (days) | 921 | 23.4 ±10.4 | 24.0 ±10.5 | MD=-0.6 (-2.1, 0.7), p=0.41 |
| Any death by day 30 (n, %) | 921 | 141 (23.0) | 63 (20.5) | RR=1.12 (0.86, 1.46), p=0.40 |
| Length of stay in ICU by day 30 (days) | 921 | 4.3 ±7.9 | 3.7 ±7.1 | MD=0.7 (-0.3, 1.7), p=0.22 |
| Need for kidney replacement therapy by day 30 (n, %) | 910<sup>c</sup> | 10 (1.6) | 6 (2.0) | RR=0.83 (0.31, 2.27), p=0.72 |
| Serious adverse event by day 30 (n, %) | 921 | 205 (33.4) | 81 (26.4) | RR=1.27 (1.02, 1.57), p=0.03 |
| Grade ≥3 events (severe) |  | 260 (42.3) | 109 (35.5) | RR=1.19 (1.00,1.42), p=0.05 |
| Grade ≥4 events (life-threatening) |  | 188 (30.6) | 74 (24.1) | RR=1.27 (1.01,1.60), p=0.04 |
| Grade 5 events (fatal) |  | 141 (23.0) | 63 (20.5) | RR=1.12 (0.86,1.46), p=0.40 |
|  |  | **Convalescent**<br>**Plasma**<br>**n=625** | **Standard of**<br>**Care**<br>**n=313** |  |
| In-hospital death by day 90 (n, %) | 938<sup>d</sup> | 156 (25.0) | 69 (22.0) | RR=1.13 (0.88, 1.45), p=0.33 |
| Time to in-hospital death by day 90 | 938 | - | - | HR=1.02 (0.76, 1.35), p=0.91 |
| Length of stay in hospital by day 90 | 938 | - | - | HR=0.91 (0.80, 1.04), p=0.18 |

<sup>a</sup> Relative Risk (RR, 95% confidence interval), Hazard Ratio (HR, 95% confidence interval) and Mean Difference (MD, with 95% CI based on robust bootstrap standard errors)
<sup>b</sup>17 patients discharged before day 30 and lost to follow-up at 30 days, and 2 withdrew consent before day 30, thus outcomes collected at day 30 (primary outcome and some other secondary outcomes for day 30) were missing.
<sup>c</sup>Excluding 11 patients on chronic kidney replacement therapy at baseline
<sup>d</sup>Excluding 2 patients that withdrew consent prior to outcome

### Secondary Efficacy Outcomes and Subgroup Analyses

Secondary outcomes for the intention-to-treat population are provided in Table 2. There were no differences in mortality or intubation or other secondary efficacy outcomes. Similarly, in the per-protocol analysis there were no differences in the secondary efficacy outcomes (eTable 4). No significant differences were observed in any of the pre-specified subgroups for both the intention-to-treat (Figure 2) and per-protocol populations (eFigure 1).

**Figure 2:**
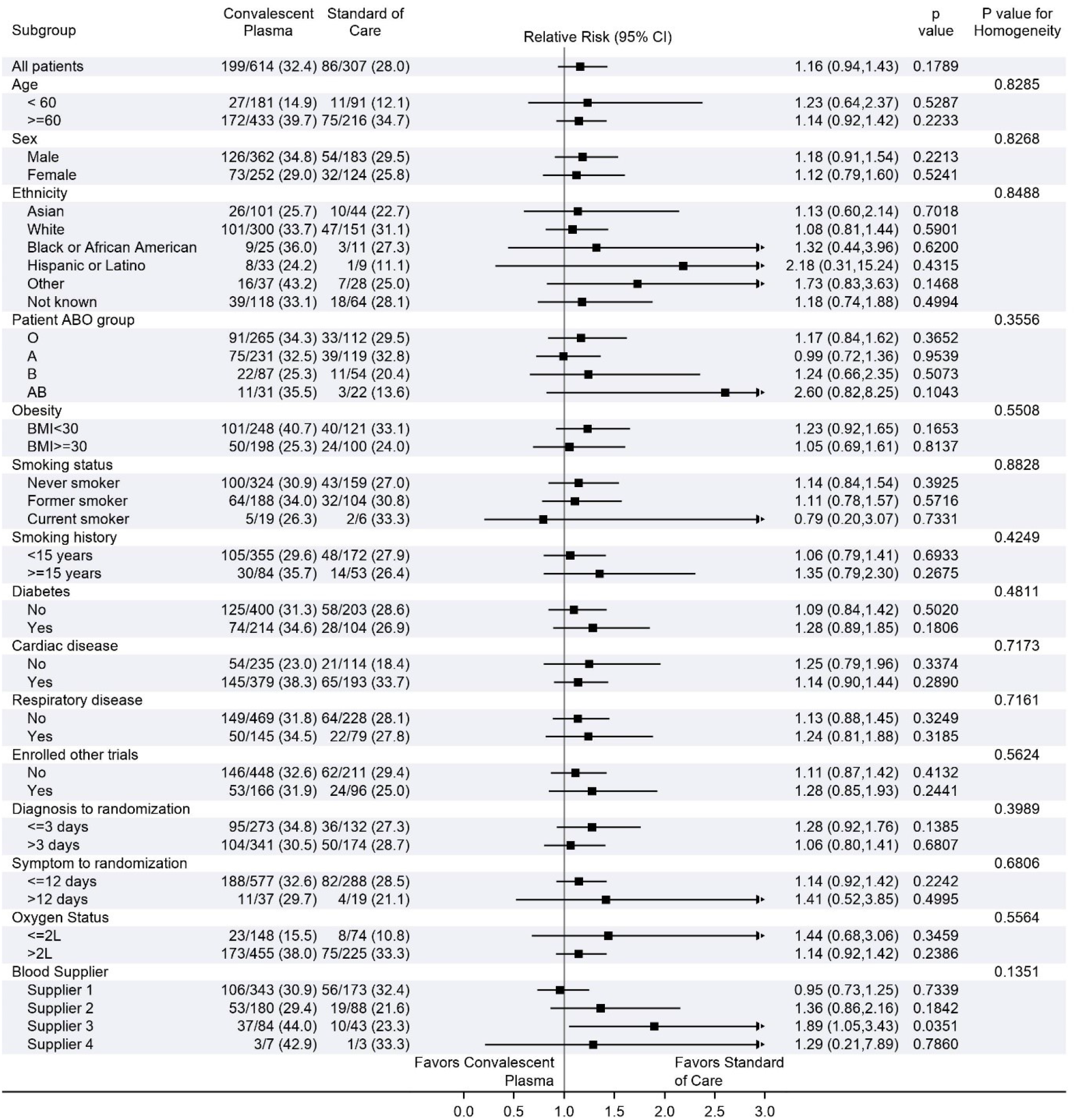
Subgroup analyses. Forest plots for the subgroup analyses for the intention to treat population. BMI: Body mass index.

### Safety

Serious adverse events occurred in 205 (33.4%) of 614 patients in the convalescent plasma arm compared to 81 (26.4%) of 307 patients in the standard of care arm for the intention-to-treat population (RR=1.27, 95% CI 1.02 to 1.57, p=0.034; eTable 5). Most of these events were worsening hypoxemia and respiratory failure. Transfusion related complications were recorded in 35 (5.7%) of 614 patients in the convalescent plasma group (eTable 7). Of the 35 reactions, 4 were life-threatening (2 transfusion-associated circulatory overload, 1 possible transfusion-related acute lung injury, 1 transfusion-associated dyspnea), and none were fatal. Thirteen of the 35 reactions were classified as transfusion-associated dyspnea. Two patients underwent serological investigation for transfusion-related acute lung injury (both negative).

### Effect-modifying role of the Antibodies in Convalescent Plasma

The distributions of antibodies in convalescent plasma units varied by blood supplier (Figure 3, eTable 9); therefore, antibody analyses controlled for supplier to address possible confounding. Transfusion of convalescent plasma with average (log-transformed) levels of ADCC yielded an odds ratio of 1.16 (95% CI: 0.85, 1.57) for the primary outcome relative to standard of care. Each one unit increase in the standardized log transformed ADCC was associated with a 24% reduction in the odds ratio of the treatment effect (OR = 0.76, 95% CI: 0.62-0.92) (Figure 3, eTable 10). This effect modifying role was also significant for the neutralization test (OR=0.77; 0.63-0.94), but not for anti-RBD ELISA (OR=0.84; 0.69-1.03), or IgG against the full transmembrane Spike (OR=1.01; 0.82-1.23).

**Figure 3:**
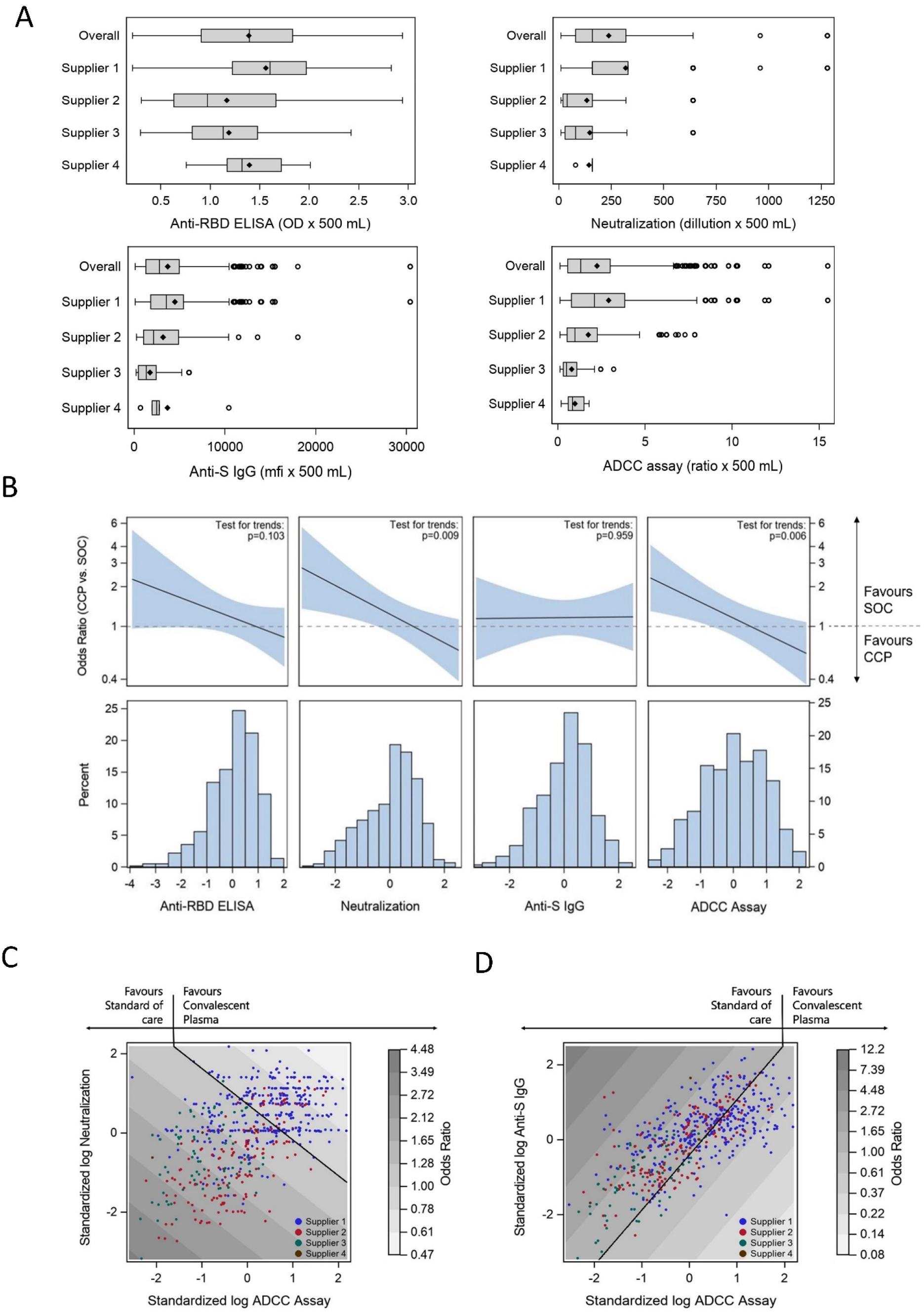
The effect-modifying role of convalescent plasma antibody content for the primary outcome. Panel A presents boxplots of absolute antibody amounts transfused to the patients in the convalescent plasma (CCP) arm for each of the serologic marker, expressed as the product of their concentration measure and the volume. Center line, median; box limits, upper and lower quartiles; whiskers, 1.5x interquartile range; points, outliers. Panel B presents the effect-modifying role of CCP antibody content for the primary outcome for each antibody measure taken individually. The top row presents the plots of the trends in the convalescent plasma effect compared to standard of care (SOC) as a function of the marker value, along with 95% confidence intervals, obtained from generalized additive models for each marker taken individually. Marker values are expressed as standard deviations of log values centered around the mean (standardized log). The horizontal dotted line represents a convalescent plasma with no effect (OR=1). The p-values imbedded in the plots (test for trend) refer to the effect modification observed with each increase of one standardized log of the marker (see table S10). The histograms in the second row present the frequency distribution of observed marker values. Panel C gives the contour plots of the odds ratio for the odds ratio for the composite event of intubation or death for individuals receiving blood as a function of the antibody-dependent cellular cytotoxicity (ADCC) ratio and the neutralization titer. Over-layed data points indicate the value of the two antibody markers for each convalescent plasma transfusion in the study, with color code indicating blood supplier. The contours are obtained from a generalized additive logistic model for the primary outcome including blood supply center, treatment and the log transformed and standardized biomarkers using smoothing splines. Panel D similarly explores the joint effect of the ADCC ratio and levels of IgG against the SARS-CoV-2 full transmembrane Spike protein on the primary outcome. ADCC: Antibody-dependent cellular cytotoxicity; CCP: COVID-19 convalescent plasma; IgG: immunoglobulin G; OR: odds ratio; RBD: receptor-binding domain; S: SARS-CoV-2 Spike protein; SOC: standard of care.

When all four serologic markers were included in the multivariate model, each one unit increase in the standardized log transformed anti-Spike IgG marker was associated with a 53% increase in the odds ratio for the deleterious effect of convalescent plasma on the primary outcome (OR=1.53, 95% CI 1.14-2.05); increases in ADCC and neutralization independently improved the effect of CCP (OR=0.66; 0.50-0.87 and OR=0.74; 0.57-0.95, respectively) while levels of anti-RBD antibodies had no effect-modifying role (OR=1.02; 0.76, 1.38) (eTable 10). There was no evidence of significant interaction between the four serologic measures in the general additive model (Figure 3, eTable 10).

### Meta-analysis

Of the fifteen other reported randomised trials, eleven used only high-titre plasma^5, 7, 22–30^ and four applied less stringent plasma selection criteria allowing for variable plasma titres^6, 31–33^. Including the results from CONCOR-1, a total of 15,301 patients were included in the trials using high-titre plasma and 968 participated in a trial applying less stringent criteria. The summary estimates for the relative risk of mortality in high-titre plasma trials was 0.97 (95% CI 0.92 – 1.02), compared to 1.25 (95% CI 0.92-1.69) in trials using unselected convalescent plasma (Figure 4).

**Figure 4.**
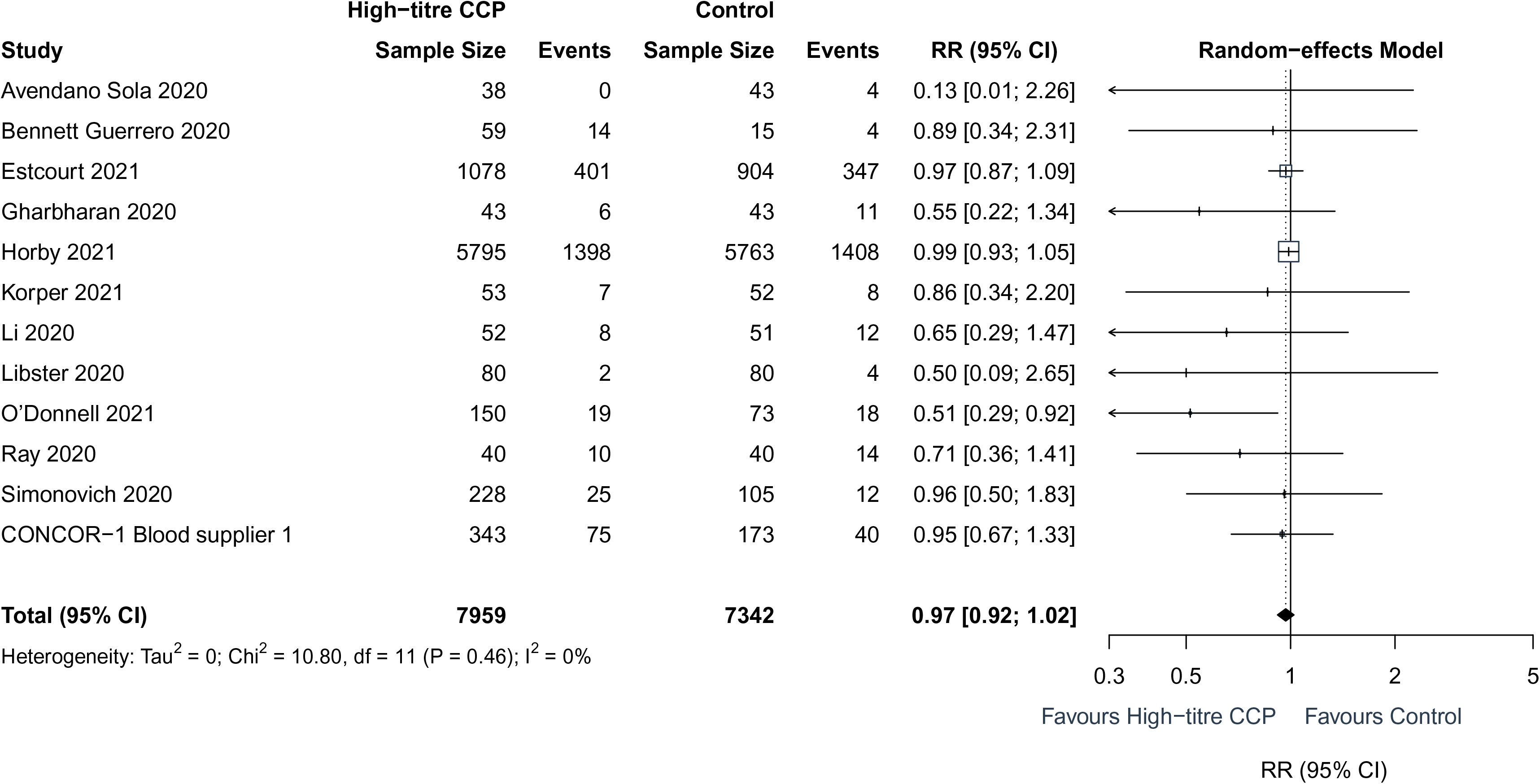
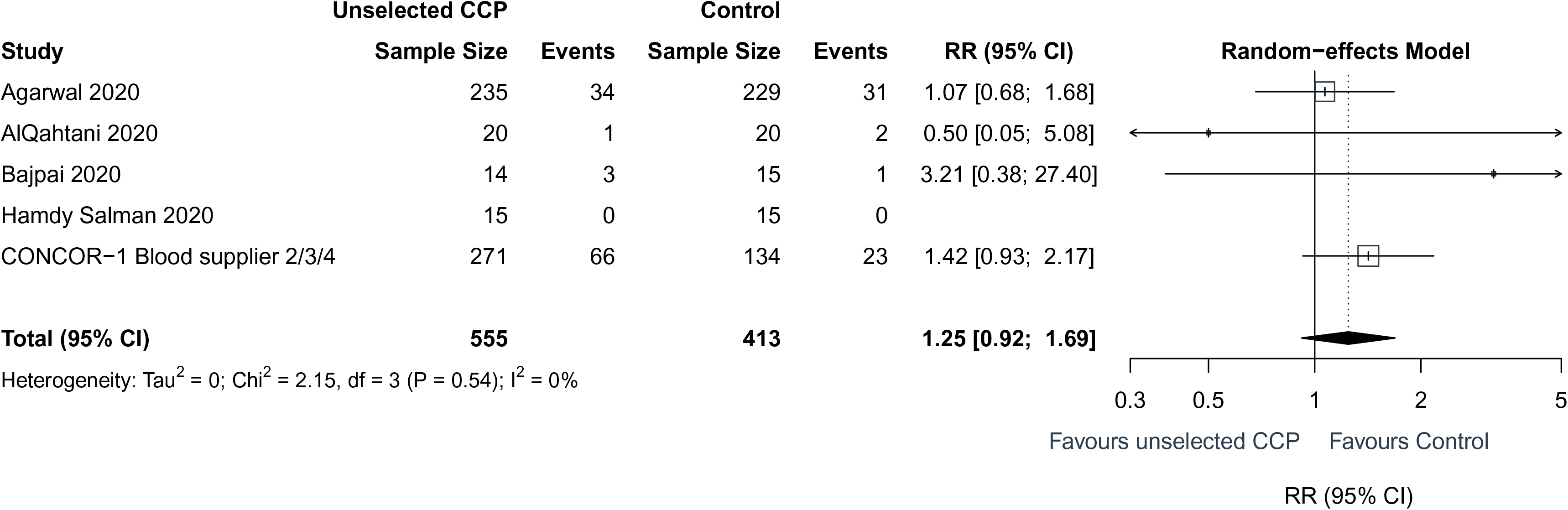
Meta-analysis of mortality at 30 days in CONCOR-1 and other trials according to convalescent plasma selection strategy. Panel A present the meta-analysis including trials that used high-titer plasma, whereas panel B presents those in which plasma which used a mix of low, medium and high-titre plasma. CCP: Covid-19 convalescent plasma; RR: relative risk.

## Discussion

The CONCOR-1 trial found that the use of convalescent plasma for the treatment of hospitalized patients with COVID-19 did not reduce the risk of intubation or death at 30 days. Patients in the convalescent plasma arm experienced more serious adverse events. Convalescent plasma was not associated with an improvement in any of the secondary efficacy outcomes or in any of the subgroups. These results are consistent with the RECOVERY trial and a recent Cochrane meta-analysis.^8, 10^ A major additional contribution of our study comes from the study of immunologic markers, which suggest that the antibody profile significantly modified the effect of convalescent plasma compared to standard of care.

The RECOVERY trial showed that transfusion of high-titer plasma was no better than standard of care in prevention of key outcomes.^10^ The US National Registry report showed that high antibody level plasma was associated with a 34% relative risk reduction in mortality compared to low antibody level plasma.^9^ Our assessment of the role of antibody profile on the clinical effect relative to standard of care is aligned with both of these conclusions. In the RECOVERY trial, plasma with a commercial ELISA cut-off corresponding to a neutralizing antibody titer of 100 or greater was used and the mortality rate ratio compared with standard of care was 1.00 (95% CI 0.93-1.07).^10^ In our trial, plasma from one of the blood suppliers (blood supplier 1) that used a similar antibody threshold (neutralizing antibody titer of 160 of greater) was associated with a similar effect size [OR 0.95 (95% CI 0.73-1.25)] (Figure 3). In contrast, the US National Registry study, which lacked a control group, reported that plasma containing high antibody levels (Ortho Vitros IgG anti-Spike subunit 1, which contains the receptor binding domain (RBD), signal to cut-off ratio > 18.45) was associated with a 34% reduction in mortality compared to plasma containing low antibody levels (signal to cut-off ratio < 4.62).^9^ In our regression model (eTable 10), plasma with anti-RBD ELISA values corresponding to this low antibody cut-off (Figure 3) would have a predicted OR of 1.49 compared with controls (95% CI: 0.98-2.29), while plasma with the corresponding high antibody cut-off would have a predicted OR of 0.91 (95% CI: 0.60-1.40), representing a 38% relative risk reduction. Thus, the 34% relative risk reduction observed by US National Registry^9^ could be explained by increased mortality with low antibody plasma, rather than improved mortality with high antibody plasma.

This conclusion is corroborated by the meta-analysis of previous trials based on plasma selection strategy. While the vast majority of patients included in convalescent plasma trials received with high-titre plasma, most patients treated outside of clinical trials did not, including many of those who received plasma by the current FDA requirements (Ortho Vitros ≥ 9.5). Only 20% of convalescent plasma included in the US National registry was considered high-titre^9^. In our study blood supplier 3 used the same plasma as the one used in clinic as part of the EUA, and in our subgroup analysis convalescent plasma from this blood supplier was associated with worse clinical outcomes (OR=1.89, 95%CI 1.05 – 3.43).

The antibody content is critical in determining the potency and potential harm of passive antibody therapy. Convalescent plasma demonstrating high levels of viral neutralization and high levels of Fc-mediated function were independently associated with a reduced risk for intubation or death. The importance of Fc-mediated function is in line with the known functional determinants of the anti-SARS-CoV-2 humoral response. In animal models of COVID-19, mutation of monoclonal antibodies leading to loss of Fc-mediated function, but sparing the neutralizing function, abrogated the protective effect of the antibody.^34–37^ In cohort studies of severe COVID-19, low Fc-mediated function but not neutralization was associated with mortality.^38, 39^

In contrast, high levels of IgG antibodies against the full transmembrane Spike protein measured by flow cytometry (which are distinct from commercial assays for IgG against Spike subunit 1) were associated with an increased risk of intubation or death after controlling for other antibody markers, suggesting that the transfusion of convalescent plasma containing non-functional anti-SARS-CoV-2 antibodies may be harmful. Antibody Fc-mediated function is dependent on the ability to aggregate and crosslink Fc receptors on target cells. This process can be disrupted by competition from other antibodies with low or absent Fc function.^40^ Similar observations were made during HIV vaccine trials, where the development of IgA antibodies against the virus envelope paradoxically increased the risk of infection due to competition with IgG,^41, 42^ and in animal models of passive immunization where transfer of antibodies could be deleterious to the host.^43^

One positive clinical trial in mild disease (n=160) found that high-titer convalescent plasma administered within 72 hours of the onset of mild COVID-19 symptoms improved clinical outcomes compared with placebo in an elderly outpatient population.^25^ Furthermore, in a Bayesian re-analysis of the RECOVERY trial, the subgroup of patients who had not yet developed anti-SARS-CoV-2 antibodies appeared to benefit from convalescent plasma.^44^ The C3PO trial, which also assessed early treatment with high-titer plasma in high-risk patients, was stopped prematurely for futility after enrolling 511 of 900 planned participants (NCT04355767). In our trial, the median time from the onset of symptoms was 8 days; however, we did not observe a difference in the primary outcome in the subgroup of patients who received convalescent plasma within three days from diagnosis.

The frequency of serious adverse events was higher in the convalescent plasma group compared with the standard of care group (33.4% vs. 26.4%; RR=1.27, 95% CI 1.02-1.57). Most of these events were caused by worsening hypoxemia and respiratory failure occurring throughout the 30 day follow up period. This frequency is consistent with the recent Cochrane review which reported an OR of 1.24 (95% CI 0.81-1.90) for serious adverse events.^8^ The frequency of transfusion-associated dyspnea and transfusion-associated circulatory overload were 2.1% and 0.8%, respectively, similar to other studies of non-convalescent plasma.^45^ The rates of transfusion reactions in CONCOR-1 were higher than what was reported in the RECOVERY trial, where transfusion reactions were reported in 13 (0.22%) of 5795 patients.^10^ CONCOR-1 site investigators included many transfusion medicine specialists and the open-label design may have encouraged reporting. However, the rate of serious transfusion-related AEs was low (4 of 614 (0.65%) convalescent plasma treated patients) and thus, does not explain the difference in serious adverse events between groups.

CONCOR-1 was a novel randomized trial designed to examine the effect of convalescent plasma versus standard of care for the primary composite outcome of intubation of death, with a capacity to explore the immunological profile of convalescent plasma and its impact on the effect of convalescent plasma. The trial involved four blood suppliers that provided local convalescent plasma units based on different antibody criteria. As a result, plasma units with a wide distribution of antibody content were included, and comprehensive antibody testing using both quantitative and functional assays provided a detailed description of the plasma product. The open-label design represents a limitation of this study as knowledge of the treatment group could influence the decision to intubate, report adverse events, or administer other treatments. The antibody profile of the recipient was unavailable at the time of this analysis. In future work we will investigate the value of convalescent plasma in patients without a detectable humoral immune response. In addition, other antibody isotypes (IgM and IgA) and IgG subclasses should be evaluated in future studies to determine their effect on clinical outcomes.

In summary, the CONCOR-1 trial did not demonstrate a difference in the frequency of intubation or death at 30 days with convalescent plasma or standard of care in hospitalized patients with COVID-19 respiratory illness. The antibody content had a significant effect-modifying role for the impact of convalescent plasma on the primary outcome. The lack of benefit and the potential concern of harm cautions against the unrestricted use of convalescent plasma for hospitalized patients with COVID-19.

## Data Availability

De-identified individual patient data with the data dictionary that underlie the reported results will be made available upon request if approved by the CONCOR-1 Steering Committee. Proposals for access should be sent to.

## Contributors

PB, JC, EJ, RCo, NMH, AT, MPZ, GBB, RB, KCL, RCa, MC, ND, DVD, DAF, AM, NR, DCS, LS, NS, AFT, RZ, AF, DMA were involved in study conceptualization, funding acquisition and methodology. PB, JC, RCo, GBB, RB, HW, AF, NL, YL and DMA developed the methodology for antibody analyses. PB, JC, NMH, AT, MPZ, GBB, LA, RB, MC, MMC, ND, DVD, JD, DAF, CG, MJG, AM, NR, BSS, DCS, LS, NS, AFT, HW, RZ, AF, DMA participated in the investigations. PB, JC, EJ, NMH, AT, MPZ, LA, RB, MMC, DVD, CG, MJG, NR, BSS and DMA were responsible for project administration. PB, JC, EJ, RC, GBB, RB, DVD, NL, YL, HW and DMA were involved in data curation. PB, JC, RC, NL, YL and DMA were responsible for formal analyses and data visualisation. PB, JC, YL, and DMA have verified the underlying data. PB, JC, RC and DMA drafted the original manuscript. All members of the writing committee reviewed and edited the final manuscript.

## Role of the funding source

Funding for the study was provided by Canadian Institutes of Health Research – COVID-19 May 2020 Rapid Research Funding Opportunity-Operating Grant; Ontario COVID-19 Rapid Research Fund; Toronto COVID-19 Action Initiative 2020 (University of Toronto); University Health Network Emergent Access Innovation Fund; University Health Academic Health Science Centre Alternative Funding Plan (Sunnybrook Health Sciences Centre); Ministère de l’Économie et de l’Innovation (Québec);  Saskatchewan Ministry of Health; University of Alberta Hospital Foundation; Alberta Health Services COVID-19 Foundation Competition; Sunnybrook Health Sciences Centre Foundation; Fondations CHU Ste-Justine; Fondation du CHUM; The Ottawa Hospital Academic Medical Organization; The Ottawa Hospital Foundation COVID-19 Research Fund; Sinai Health System Foundation and McMaster University. These did not have any role in the writing of the manuscript or the decision to submit it for publication. The authors did not receive payments from any pharmaceutical company or other agency to write this article. All the authors had full access to the full data in the study and accept responsibility to submit for publication.

## Declaration of Interests

PB reports payments to his institution from Canadian Institutes of Health Research, Ministère de l’économie et de l’innovation du Québec, Fondation du CHU Sainte-Justine , Fondation du CHUM, and Fonds de Recherche du Québec en Santé; JC reports research grants from Canadian Institutes of Health Research, University of Toronto, Sunnybrook Health Sciences Centre, and Canadian Blood Services; NMH reports research project funding unrelated to this study from Canadian Institutes of Health Research and Canadian Blood Services, honorarium for educational events unrelated to this study from CSL Behring Canada, and participation on a Data Safety Monitoring Board or Advisory Board for Canadian Blood Services (Scientific Advisory Committee); MPZ reports consulting fees from Canadian Blood Services in her role as a Medical Officer but receives no direct benefit from the study results; GBB reports a grant from Canadian Institutes of Health Research for Masters students; MMC reports consulting fees from Octapharma, participation in a data safety monitoring board or advisory board for Cerus and Haemonetics, and receipt of equipment, materials, drugs, medical writing, gifts or other services by her institution from Cerus; ND reports grants from AFP Innovation Fund Award Sunnybrook Health Sciences Centre, and University of Toronto; MJG reports payments to his institution from Regeneron, consulting fees from Regeneron and ReAlta Life Sciences, and participation on a Data  Safety Monitoring Board or Advisory Board  for Enzychem and Sobi; BSS reports employment by New York Blood Centre (but receiving no direct benefit from study participation); LS reports a leadership or fiduciary role in other board, society, committee or advocacy group, paid or unpaid (Member of the Ontario Bioethics Table for COVID-19); NS reports payment to her institution from the MSH Foundation for CONCOR;  AF reports funding from Fondation du CHUM, Ministère de l’économie et de l’Innovation du Québec, and  Canadian Institutes of Health Research. All other authors declare no competing interests.

## Data Sharing

De-identified individual patient data with the data dictionary that underlie the reported results will be made available upon request if approved by the CONCOR-1 Steering Committee. Proposals for access should be sent to. The protocol and statistical analysis plan are available online.

## CONCOR-1 Online-Only Appendix

### List of CONCOR-1 Investigators

**Principal investigators**: Donald Arnold MD, Philippe Bégin MD PhD, Jeannie Callum MD

**Writing Committee:** Donald Arnold MD, Philippe Bégin MD PhD, Jeannie Callum MD, Erin Jamula MSc, Nancy Heddle MSc, Richard Cook PhD, Andrés Finzi PhD, Luiz Amorim MD, Guillaume Beaudoin-Bussières PhD, Renée Bazin PhD, Richard Carl, Michaël Chassé MD PhD, PhD, Melissa M. Cushing MD, Nick Daneman MD, Dana V. Devine PhD, Jeannot Dumaresq MD, Dean A. Ferguson PhD, Marshall Glesby MD, PhD, Na Li PhD, Yang Liu MMath, Kent Cadogan Loftsgard, Alison McGeer MD, Nancy Robitaille MD, Bruce S. Sachais MD, PhD, Damon C. Scales MD PhD, Lisa Schwartz PhD, Nadine Shehata MD, Alan Tinmouth MD, Alexis F. Turgeon MD MSc, Heidi Wood PhD, Ryan Zarychanski MD MSc, Michelle Zeller MD

**Steering Committee**: Luiz Amorim MD, Renée Bazin PhD, Richard Carl, Michaël Chassé MD PhD, Richard Cook PhD, Melissa M. Cushing MD, Nick Daneman MD, Dana V. Devine PhD, Dean A. Ferguson PhD, Nancy Heddle MSc, Kent Cadogan Loftsgard, Alison McGeer MD, Nancy Robitaille MD, Bruce S. Sachais MD PhD, Damon C. Scales MD PhD, Lisa Schwartz PhD, Nadine Shehata MD, Alan Tinmouth MD, Alexis F. Turgeon MD MSc, Ryan Zarychanski MD MSc, Michelle Zeller MD

**Central Methods Centre:** Julie Carruthers, Erin Jamula, Kayla Lucier; McMaster Centre for Transfusion Research

**Québec Study Coordination**: Marie-Christine Auclair, Meda Avram, Michael Brassard, Sabrina Cerro, Véronica Martinez, Julie Morin, Marie Saint-Jacques, Maxime Veillette

**Logistics Methods Centre:** Chantal Armali, Amie Kron, Dimpy Modi; University of Toronto Quality in Utilization, Education and Safety in Transfusion (QUEST) Research Program, Sunnybrook Health Sciences Centre

**Database Design and Management**: Joanne Duncan, Pauline Justumus, Melanie St John, Geneviève St-Onge, Milena Hadzi-Tosev

**Study Monitors:** Jackie Amaral, Tanja Cerovina, Mila Khanna, Monika Wiseman, Karlee Trafford, Samantha Libfeld, Tamara Bright, Louise Rousseau, Rocco Paniccia, Sergio Assis, Daniele Aguiar; Ozmosis Research Inc.

**Community Advisory Committee**: Pierre-Marc Dion, Kent Cadogan Loftsgard*, Lawrence McGillivary, Andre Valleteau de Moulliac, Sheila A. Nyman, Stephanie Perilli, Paulette Jean Van Vliet, Nancy Heddle, Shannon Lane, Katerina Pavenski, Rebecca Pereira, Emily Sirotich (Advisors: Julie Abelson, Saara Greene, Lisa Schwartz) *referred to CONCOR-1 research partnership recruitment by the coordinators of the Strategy for Patient-Oriented Research – Primary and Integrated Health Care Innovations Network. - https://spor-pihci.com

**Communications Committee:** Michelle Zeller (Chair), Kayla Lucier, Aditi Khandelwal, Adrienne Silver, Dana Ellingham, Joanne Duncan, Caroline Gabe, Anne Trueman, Ana Catalina Alvarez Elias,Natasha Jawa, Julia Upton, Laurent Paul Ménard, Chantal Armali, Florence Meney, Rulan Parekh, Eric Dimitri, Menaka Pai, Julie Carruthers

**Inventory and Distribution Committee:** Alan Tinmouth (Chair), Michelle Zeller, Melanie St. John, Swarni Thakar, Sarah Longo

**Antibody-dependent cellular cytotoxicity assay and flow cytometry:** Centre Hospitalier de l’Université de Montréal Research Center: Guillaume Beaudoin-Bussières, Sai Priya Anand, Mehdi Benlarbi, Catherine Bourassa, Marianne Boutin, Jade Descôteaux-Dinelle, Gabrielle Gendron-Lepage, Guillaume Goyette, Annemarie Laumaea, Halima Medjahed, Jérémie Prévost, Jonathan Richard, Andrés Finzi.

**Virus neutralization assay:** Zoonotic Diseases and Special Pathogens, National Microbiology Laboratory, Public Health Agency of Canada, Winnipeg, Manitoba: Michael Drebot, Heidi Wood, Alyssia Robinson, Emelissa Mendoza, Kristina Dimitrova, Kathy Manguiat, Clark Phillipson, Michael Chan; Medical Microbiology & Immunology, University of Alberta, Edmonton, Alberta: David Evans, James Lin.

**Anti-RBD antibody:** Héma-Québec : Lucie Boyer, Marc Cloutier, Mathieu Drouin, Éric Ducas, Nathalie Dussault, Marie-Josée Fournier, Patricia Landy, Marie-Ève Nolin, Josée Perreault, Tony Tremblay.

**Health Economics Expertise:** Feng Xie PhD

Participating Sites:

**British Columbia**

Abbotsford Regional Hospital, Abbotsford, British Columbia: Matthew Yan (Principal Investigator), David Liu, Michelle Wong, Gus Silverio, Kristin Walkus, Mikaela Barton, Katherine Haveman, Darlene Mueller, Ashley Scott

Royal Jubilee Hospital & Victoria General Hospital, Victoria, British Columbia: Daniel Ovakim (Principal Investigator), Matthew Moher, Gordon Wood, Tracey Roarty, Fiona Auld, Gayle Carney, Virginia Thomson

St. Paul’s Hospital, Vancouver, British Columbia: David Harris (Principal Investigator), Rodrigo Onell, Keith Walley, Katie Donohoe, Crystal Brunk, Geraldine Hernandez, Tina Jacobucci, Lynda Lazosky, Puneet Mann, Geeta Raval, Ligia Araujo Zampieri

Vancouver General Hospital, Vancouver, British Columbia: Andrew Shih (Principal Investigator), Mypinder Sekhon, Alissa Wright, Nicola James, Gaby Chang, Roy Chen, Kanwal Deol, Jorell Gantioqui, Elyse Larsen, Namita Ramdin, Margaret Roche, Kristin Rosinski, Lawrence Sham, Michelle Storms

**Alberta**

Foothills Hospital, Peter Lougheed Centre, Rockyview General Hospital, Calgary, Alberta: Davinder Sidhu (Principal Investigator), Mark Gillrie, Etienne Mahe, Deepa Suryanarayan, Alejandra Ugarte-Torres, Traci Robinson, Mitchell Gibbs, Julia Hewsgirard, Marnie Holmes, Joanna McCarthy, Meagan Ody

University of Alberta, Royal Alexandra Hospital, Edmonton, Alberta: Susan Nahirniak (Principal Investigator), Karen Doucette, Wendy Sligl, Ashlesah Sonpar, Kimberley Robertson, Jeffrey Narayan, Leka Ravindran, Breanne Stewart, Lori Zapernick

**Saskatchewan**

Pasqua Hospital, Regina General Hospital, Regina, Saskatchewan: Donna Ledingham (Principal Investigator), Stephen Lee, Eric Sy, Alexander Wong, Karolina Gryzb, Sarah Craddock, Dennaye Fuchs, Danielle Myrah, Sana Sunny

Royal University Hospital, St. Paul’s Hospital, Saskatoon, Saskatchewan: Oksana Prokopchuk-Gauk (Principal Investigator), Sheila Rutledge Harding, Siddarth Kogilwaimath, Nancy Hodgson, Dawn Johnson, Simona Meier, Kim Thomson

**Manitoba**

Grace General Hospital, Health Sciences Centre, St. Boniface General Hospital, Winnipeg, Manitoba: Arjuna Ponnampalam (Principal Investigator), Emily Rimmer (Principal Investigator), Amila Heendeniya (Principal Investigator), Brett Houston, Yoav Kenyan, Sylvain Lother, Kendiss Olafson, Barret Rush, Terry Wuerz, Ryan Zarychanski, Dayna Solvason, Lisa Albensi, Soumya Alias, Nora Choi, Laura Curtis, Maureen Hutmacher, Hessam Kashani, Debra Lane, Nicole Marten, Tracey Pronyk-Ward (Canadian Blood Services), Lisa Rigaux, Rhonda Silva, Quinn Tays

**Ontario**

Bluewater Health, Sarnia, Ontario: Glenna Cuccarolo (Principal Investigator), Renuka Naidu, Jane Mathews, Margaret Mai, Victoria Miceli, Liz Molson, Gayathri Radhakrishnan, Linda Schaefer, Michel Haddad, Shannon Landry

Grand River Hospital and St. Mary’s General Hospital, Kitchener, Ontario: Colin Yee (Principal Investigator), Robert Chernish, Rebecca Kruisselbrink, Theresa Liu, Jayna Jeromin, Atif Siddiqui, Carla Girolametto, Kristin Krokoszynski,

Hamilton General Hospital, Hamilton, Ontario: Menaka Pai (Principal Investigator), Daniela Leto, Cheryl Main, Alison Fox-Robichaud, Bram Rochwerg, Michelle Zeller, Erjona Kruja, Dana Ellingham, Erin Jamula, Meera Karunakaran, Shannon Lane, Kayla Lucier, Disha Sampat, Ngan Tang

Juravinski Hospital, Hamilton, Ontario: Michelle Zeller (Principal Investigator), Daniela Leto, Bram Rochwerg, Erjona Kruja, Dana Ellingham, Erin Jamula, Meera Karunakaran, Shannon Lane, Kayla Lucier, Disha Sampat, Ngan Tang

Lakeridge Health, Oshawa and Ajax, Ontario: Karim Soliman (Principal Investigator), Daniel Ricciuto, Kelly Fusco, Taneera Ghate, Holly Robinson

London Health Sciences Centre, London, Ontario: Ziad Solh (Principal Investigator), Ian Ball, Sarah Shalhoub, Marat Slessarev, Michael Silverman, Eni Nano, Tracey Bentall, Eileen Campbell, Jeffery Kinney, Seema Parvathy, and the Medical Laboratory Technologists at London Health Sciences Centre

Markham Stouffville Hospital, Markham, Ontario: Valerie Sales (Principal Investigator), Evridiki Fera, Anthony La Delfa, Jeya Nadarajah, Henry Solow, Edeliza Mendoza, Katrina Engel, Diana Monaco, Laura Kononow, Sutharsan Suntharalingam

Mount Sinai Hospital, Toronto, Ontario: Nadine Shehata (Principal Investigator), Mike Fralick, Allison McGeer, Laveena Munshi, Samia Saeed, Omar Hajjaj, Elaine Hsu, and the Medical Laboratory Technologists at Mount Sinai Hospital

Niagara Health System, St. Catharines Site, St. Catharines, Ontario: Jennifer LY Tsang (Principal Investigator), Karim Ali, Erick Duan, George Farjou, Lorraine Jenson, Mary Salib, Lisa Patterson, Swati Anant, Josephine Ding, Jane Jomy

North York General Hospital, North York, Ontario: Eneko Arhanchiague (Principal Investigator), Pavani Das, Anna Geagea, Sarah Ingber, Elliot Owen, Alexandra Lostun, Tashea Albano, Antara Chatterjee, Manuel Giraldo, Jennifer Hickey, Ida Lee, Nea Okada, Nicholas Pasquale, Romina Ponzielli, Mary Rahmat, Shelina Sabur, Maria Schlag, Leonita Aguiar, Ashmina Damani, Suhyoung Hong, Mona Kokabi, Carolyn Perkins

The Ottawa Hospital, Ottawa, Onatrio: Alan Tinmouth (Principal Investigator), Juthaporn Cowan, Tony Giulivi, Derek MacFadden, Holly Carr (lead RC), Joe Cyr, Amanda Pecarskie, Rebecca Porteous, Priscila Ogawa Vedder, Irene Watpool.

Queensway Carleton Hospital, Ottawa, Ontario: Moira Rushton-Marovac (Principal Investigator), Phil Berardi, Laith Bustani, Alison Graver, Akshai Iyengar, Magdalena Kisilewicz, Jake Majewski, Misha Marovac, Ruchi Murthy, Karan Sharma, Marina Walcer

St. Joseph’s Healthcare, Hamilton, Ontario: Shuoyan Ning (Principal Investigator), Zain Chagla, Jason Cheung, Erick Duan, France Clarke, Karlo Matic, Manuel Giraldo, Jennifer Hickey, Ida Lee, Nea Okada, Nicholas Pasquale, Romina Ponzielli, Mary Rahmat, Shelina Sabur, Maria Schlag

St. Joseph’s Health Centre, Toronto, Ontario: Katerina Pavenski (Principal Investigator), Travis Carpenter, Kevin Schwartz, Paril Suthar, Aziz Jiwajee, Daniel Lindsay, Aftab Malik, Brandon Tse

St. Michael’s Hospital, Toronto, Ontario: Katerina Pavenski (Principal Investigator), Larissa Matukas, Joel Ray, Paril Suthar, Shirley Bell, Aziz Jiwajee, Elizabeth Krok, Daniel Lindsay, Aftab Malik, Brandon Tse

Scarborough Health Network, Scarborough, Ontario: Rosemarie Lall (Principal Investigator), Ray Guo, Susan John, Vishal Joshi, Jessica Keen, Chris Lazongas, Jacqueline Ostro, Kevin Shore, Jianmin Wang, Jincheol Choi, Pujitha Nallapati, Tina Irwin, Victor Wang, Petra Sheldrake

Sunnybrook Health Sciences Centre, Toronto, Ontario: Yulia Lin (Principal Investigator), Neill Adhikari, Jeannie Callum, Nick Daneman, Hannah Wunsch, Amie Kron, Chantal Armali, Jacob Bailey, Harley Meirovich, Dimpy Modi, Connie Colavecchia

Trillium Health Partners, Mississauga, Ontario: Christopher Graham (Principal Investigator), Eiad Kahwash, Sachin Sud, Martin Romano

University Health Network, Toronto, Ontario: Christine Cserti-Gazdewich (Principal Investigator), Bryan Coburn, Lorenzo Del Sorbo, John Granton, Shahid Husain, Jacob Pendergrast, Abdu Sharkawy, Liz Wilcox, Samia Saeed, Chantal Armali, Omar Hajjaj, Maria Kulikova, Sophia Massin

Windsor Regional Hospital, Windsor, Ontario: Caroline Hamm (Principal Investigator), Wendy Kennette, Ian Mazzetti, Krista Naccarato, Grace Park, Alex Pennetti, Corrin Primeau, Cathy Vilag

**Québec**

Centre Hospitalier de l’Université de Montréal (CHUM), Montréal, Québec: Madeleine Durand (Principal Investigator), Michaël Chassé, Yves Lapointe, Anne-Sophie Lemay, Emmanuelle Duceppe, Benjamin Rioux-Massé, Cécile Tremblay, Pascale Arlotto, Claudia Bouchard, Stephanie Matte, Marc Messier-Peet, COVID-19 Unit Personnel

CHU de Québec-Université Laval, Québec city, Québec: Alexis F. Turgeon (Principal Investigator), Charles-Langis Francoeur, François Lauzier, Vincent Laroche, Guillaume Leblanc, David Bellemare, Ève Cloutier, Olivier Costerousse, Émilie Couillard Chénard, Rana Daher, Marjorie Daigle, Stéphanie Grenier, Gabrielle Guilbeault, Marie-Pier Rioux, Maude St-Onge, Antoine Tremblay

CHU de Sherbrooke, Sherbrooke, Québec: Alexandra Langlois (Principal Investigator), Brian Beaudoin, Luc Lanthier, Pierre Larrivée, Pierre-Aurèle Morin, Élaine Carbonneau, Robert Lacasse

CHU Sainte-Justine, Montréal, Québec: Olivier Drouin (Principal Investigator), Julie Autmizguine, Philippe Bégin, Isabelle Boucoiran, Geneviève Du Pont Thibodeau, Meda Avram, Mary-Ellen French, Annie La Haye, Vincent Lague, Karine Léveillé, Nancy Robitaille

CISSS Montérégie-Centre, Hôpital Charles-Lemoyne, Greenfield Park, Québec: Nadim Srour (Principal Investigator), Susan Fox, Diaraye Baldé, Lorraine Ménard, Suzanne Morissette, Miriam Schnorr-Meloche, Andrée-Anne Turcotte, Caroline Vallée

Hôpital Cité-de-la-Santé de Laval, Laval, Québec: Danielle Talbot (Principal Investigator), Stéphanie Castonguay, Tuyen Nguyen, Natalie Rivest, Marios Roussos, Esther Simoneau, Andreea Belecciu, Marie-Hélène Bouchard, Eric Daviau and his team, Cynthia Martin, Nicole Sabourin, Solange Tremblay

Hôpital de Chicoutimi, Chicoutimi, Québec: François Ménard (Principal Investigator), Émilie Gagné, Nancy-Lisa Gagné, Julie Larouche, Vanessa Larouche, Véronick Tremblay, Vicky Tremblay

Hôpital de Trois-Rivières, Trois-Rivières, Québec: André Poirier (Principal Investigator), Pierre Blanchette, David Claveau, Marianne Lamarre, Danielle Tapps

Hôpital du Sacré-Cœur-de-Montréal, Montréal, Québec: Christine Arseneault (Principal Investigator), Martin Albert, Anatolie Duca, Jean-Michel Leduc, Annie Barsalou, Suzanne Deschênes-Dion, Stéphanie Ibrahim, Stéphanie Ridyard, Julie Rousseau

Hôpital Maisonneuve-Rosemont, Montréal, Québec: Mélissa Boileau (Principal Investigator), Stéphane Ahern, Marie-Pier Arsenault, Simon-Frédéric Dufresne, Luigina Mollica, Hang Ting Wang, Soizic Beau, Dominique Beaupré, Marjolaine Dégarie, Iris Delorme, Melissa Farkas, Michel-Olivier Gratton, Arnaud Guertin, Guylaine Jalbert, Mélanie Meilleur, Charles Ratté Labrecque, Élaine Santos, Julie Trinh Lu

Hôpital régional de St-Jérôme, St-Jérôme, Québec: Sébastien Poulin (Principal Investigator), Julien Auger, Marie-Claude Lessard, Louay Mardini, Yves Pesant, Laurie Delves, Lisa Delves, Sophie Denault, Sofia Grigorova, Michelle Lambert, Nathalie Langille, Corinne Langlois, Caroline Rock, Yannick Sardin Laframboise

Hôtel-Dieu de Lévis, Lévis, Québec: Jeannot Dumaresq (Principal Investigator), Danièle Marceau, Patrick Archambault, Joannie Bélanger Pelletier, Estel Duquet-Deblois, Vanessa Dupuis-Picard, Yannick Hamelin, Samuel Leduc, Mélanie Richard

Institut de Cardiologie et Pneumologie de Québec – Université Laval, Québec city, Québec: Andréanne Côté (Principal Investigator), Marc Fortin, Philippe Gervais, Vincent Laroche, Marie-Ève Boulay, Claudine Ferland, Jakie Guertin, Johane Lepage, Annie Roy, BB and COVID-19 unit personnel

Jewish General Hospital, Montréal, Québec: Christina Greenaway (Principal Investigator), Sarit Assouline, Stephen Caplan, Ling Kong, Christina Canticas, Carley Mayhew, Johanne Ouedraogo, Tévy-Suzy Tep

McGill University Hospital Center, Montréal, Québec: Makeda Semret (Principal Investigator), Matthew Cheng, Marina Klein, Nadine Kronfli, Patricia Pelletier, Salman Qureshi, Donald Vinh, Robert Dziarmaga, Hansi Peiris, Karène Proulx-Boucher, Jonathan Roger, Molly-Ann Rothschild, Chung-Yan Yuen

Hôtel-Dieu de Lévis, Lévis, Québec: Jeannot Dumaresq (Principal Investigator), Patrick Archambault, Danièle Marceau, Joannie Bélanger Pelletier, Estel Duquet-Deblois, Vanessa Dupuis-Picard, Yannick Hamelin, Samuel Leduc, Mélanie Richard

**New Brunswick**

Vitalité Health Network, Moncton, New Brunswick: Gabriel Girouard (Principal Investigator), Richard Garceau, Rémi LeBlanc, Eve St-Hilaire, Patrick Thibeault, Karine Morin, Gilberte Caissie, Jackie Caissie Collette, Line Daigle, Mélissa Daigle, Bianca Gendron, Nathalie Godin, Angela Lapointe, Gabrielle Moreau, Lola Ouellette-Bernier, Joanne Rockburn, Brigitte Sonier-Ferguson, Christine Wilson, and the many collaborating nursing staff on the ICU and COVID units.

**New York**

Weill Cornell Medicine, New York City, New York: Marshall Glesby (Principal Investigator), Melissa M. Cushing, Robert DeSimone, Grant Ellsworth, Rebecca Fry, Noah Goss, Roy Gulick, Carlos Vaamonde, Timothy Wilkin, Celine Arar, Jonathan Berardi, Dennis Chen, Cristina Garcia-Miller, Arthur Goldbach, Lauren Gripp, Danielle Hayden, Kathleen Kane, Jiamin Li, Kinge-Ann Marcelin, Christina Megill, Meredith Nelson, Ailema Paguntalan, Gabriel Raab, Gianna Resso, Roxanne Rosario, Noah Rossen, Shoran Tamura, Ethan Zhao

New York-Presbyterian Brooklyn Methodist Hospital, New York City, New York: Andy Huang (Principal Investigator), Cheryl Goss, Young Kim, Eshan Patel, Sonal Paul, Tiffany Romero, Naima ElBadri, Lina Flores, Tricia Sandoval

New York-Presbyterian Lower Manhattan Hospital, New York City, New York: Harjot Singh (Principal Investigator), Shashi Kapadia, Ljiljana Vasovic, Shanna-Kay Griffiths, Daniel Alvarado, Fiona Goudy, Melissa Lewis, Marina Loizou, Rita Louie

**Brazil**

Hemorio, Hospital and Regional Blood Center, Rio de Janeiro: Luiz Amorim Filho (Principal Investigator), Rodrigo Guimaraes, Maria Esther Lopes, Margarida Pêcego, Caroline Gabe, Natalia Rosario, Carlos Alexandre da Costa Silva, Thais Oliveira, Maria Cristina Lopes, Sheila Mateos

**Blood Suppliers**

Canadian Blood Services: Dana Devine, Chantale Pambrun, Sylvia Torrance, Steven Drews, Janet McManus, Oriela Cuevas, Wanda Lafresne, Patrizia Ruoso, Christine Shin, Tony Steed, Rachel Ward, and the many CBS staff who assisted with the collection and provision of COVID-19 convalescent plasma.

Héma-Québec: Isabelle Allard, Renée Bazin, Marc Germain, Sébastien Girard, Éric Parent, Claudia-Mireille Pigeon, Nancy Robitaille.

Hemorio, Hospital and Regional Blood Center, Rio de Janeiro: Luiz Amorim Filho (Principal Investigator), Rodrigo Guimaraes, Maria Esther Lopes, Margarida Pêcego, Caroline Gabe, Natalia Rosario, Carlos Alexandre da Costa Silva, Thais Oliveira, Maria Cristina Lopes, Sheila Mateos

New York Blood Center: Lucette Hall, Sarai Paradiso, Bruce Sachais, Donna Strauss

The authors would like to acknowledge recovered patients that donated their plasma, the staff involved in the recruitment, collection, qualification, and distribution of COVID-19 convalescent plasma and the staff of the COVID-19 wards who made the study possible.

The authors would also like to thank the members of the **Independent Data Safety Monitoring Committee**: Keyvan Karkouti MD, Robert Fowler MD, Meghan Delaney MD, George Tomlinson PhD, Darryl Davis MD, and Boris Juelg MD.

### Supplemental methods

#### Definitions

##### Adverse event (AE)

An AE is any untoward medical occurrence that occurs in a patient or clinical investigation subject administered a pharmaceutical product, and which does not necessarily have a causal relationship with the treatment assignment. An AE can therefore be any unfavorable and unintended sign (including abnormal laboratory finding), symptom, or disease temporally associated with the use of an investigational product, whether or not considered related to the product.

##### Adverse drug reaction (ADR)

An ADR is any noxious and unintended response to an IMP related to any dose. The phrase ‘response to an IMP’ means that a causal relationship between the IMP and an AE carries at least a reasonable possibility, i.e., the relationship cannot be ruled out.

##### Other significant AEs

Any marked laboratory abnormalities or any AEs that lead to an intervention, including withdrawal of drug treatment, dose reduction, or significant additional concomitant therapy.

##### Severity of AEs

The severity of each AE must be assessed by the Investigator and graded based on CTCAE v4.03. A clinical AE NOT identified elsewhere in the grading table will be graded using one of the following categories:

1. **Mild**: Mild symptoms causing no or minimal interference with usual social & functional activities with intervention not indicated

2. **Moderate**: Moderate symptoms causing greater than minimal interference with usual social & functional activities with intervention indicated

3. **Severe**: Severe symptoms causing inability to perform usual social and functional activities with intervention or hospitalization indicated

4. **Life-threatening:** Potentially life-threatening symptoms causing inability to perform basic self-care functions with intervention indicated to prevent permanent impairment, persistent disability, or death.

5. **Death:** The AE resulted in the subject’s death.

##### Serious AE (SAE)

A SAE is generally defined as any untoward medical occurrence that at any dose:

1. Results in death;

2. Is life-threatening; this means that the subject is at risk of death at the time of the event; it does not mean that the event hypothetically might have caused death if it was more severe;

3. Requires hospitalization (overnight or longer) or prolongation of existing hospitalization or invasive procedure;

4. Results in persistent or significant disability or incapacity;

5. Results in congenital anomaly or birth defect;

6. Is not be immediately life-threatening or result in death or hospitalization but may jeopardize the subject or require intervention to prevent one of the above outcomes.

#### Inclusion/Exclusion Criteria

##### Inclusion criteria

1. ≥16 years of age (≥18 years of age in the United States, Brazil, and Israel)
2. Admitted to hospital for confirmed COVID-19 respiratory illness
3. Receiving supplemental oxygen
4. 500 mL of ABO compatible CCP is available

##### Exclusion criteria

1. Onset of signs or symptoms of COVID-19 respiratory illness >12 days prior to randomization (eg. cough, chest pain, dyspnea, or hypoxia)
2. Intubated or plan in place for intubation
3. Plasma is contraindicated (e.g. history of anaphylaxis from transfusion)
4. Decision in place for no active treatment

### Study outcomes

#### Primary outcomes

The primary outcome is a composite of intubation or death at Day 30.

#### Secondary outcomes

- Time to intubation or death
- Ventilator-free days at day 30
- In-hospital death by Day 90
- Time to in-hospital death
- Death by Day 30
- Length of stay in ICU
- Length of stay in hospital
- Need for ECMO
- Need for renal replacement therapy
- Myocarditis
- Patient-reported outcome at Day 30
- Incremental cost per quality-adjusted life year (QALY)
- CPP transfusion-associated AEs
- Grade 3 and 4 SAEs
- Cumulative incidence of Grade 3 and 4 SAEs (CTCAE)

### Electronic Case Report Form

Study data were collected and managed using REDCap electronic data capture tools hosted at McMaster University. REDCap (Research Electronic Data Capture) is a secure, web-based application designed to support data capture for research studies, providing: 1) an intuitive interface for validated data entry; 2) audit trails for tracking data manipulation and export procedures; 3) automated export procedures for seamless data downloads to common statistical packages; and 4) procedures for importing data from external sources.^1^

### Antibody assays

#### Anti-RBD ELISA

Recombinant SARS-CoV-2 S RBD proteins (2.5 μg/ml), were prepared in PBS and were adsorbed to plates (100 µl/well; Immulon 2HB, Thermo Scientific) overnight at 4°C. Coated wells were subsequently blocked with blocking buffer (Phosphate-buffered saline [PBS] containing 0.1% Tween20 and 2% BSA) for 1h at room temperature. Wells were then washed four times with washing buffer (PBS containing 0.1% Tween20). CR3022 mAb (50ng/ml final concentration) or plasma from SARS-CoV-2-infected or uninfected donors (1/100 dilution)in blocking buffer) were incubated with the RBD-coated wells for 1h at room temperature. Plates were washed four times with washing buffer followed by incubation with HRP conjugated goat anti-human IgA+IgG+IgM diluted in blocking buffer (Jackson ImmunoResearch Laboratories, Inc.) for 1h at room temperature, followed by four washes. HRP enzyme activity was determined after a 20 minute incubation with 3,3’,5,5’-Tetramethylbenzidine (TMB, ESBE Scientific) followed by blocking with H2SO4 1N. Plates were read using a microplate reader (Synergy H1, Bio-Tek) within 30 min after blocking the reaction. The cut-off value for seropositivity, set at 0.250, was calculated using the mean OD + 3 standard deviations of 13 COVID-19 negative plasma samples (collected in 2019, before the outbreak of SARS-CoV-2) plus a 15% inter-assay coefficient of variation. Following implementation of the assay, we used results from the analysis of 94 convalescent plasma samples and 88 negative samples and the determined cut-off value yielded a sensitivity of 98.9% and a specificity of 98.5%.

#### Cell surface staining

293T and 293T.SARS-CoV-2.Spike cells^2^ were mixed at a 1:1 ratio and stained with the anti-RBD monoclonal antibody CR3022 (5 µg/mL) or plasma (1/250 dilution) for 45 minutes at 37°C. Cells were then washed 2 times with PBS before being incubated with secondary antibodies. AlexaFluor-647 conjugated goat anti-human IgA, IgM, IgM+IgG+IgA (1,70 µg/mL; Jackson ImmunoResearch) or AlexaFluor-647 conjugated mouse anti-human IgG (3 µg/mL; Biolegend) were used as secondary antibodies to stain cells for 20 minutes at room temperature. Cells were then washed 2 times with PBS before being fixed in 2 % PBS-formaldehyde solution. The percentage of transduced cells (GFP+ cells) was determined by gating on the living cell population based on viability dye staining (AquaVivid, Thermo Fisher Scientific). Samples were acquired on a LSRII cytometer (BD Biosciences) and data analysis was performed using FlowJo v10.7.1 (Tree Star).

#### Antibody-dependent cellular cytotoxicity (ADCC) assay

ADCC was measured as described previously.^3^ Briefly, parental CEM.NKr CCR5+ cells were mixed at a 1:1 ratio with CEM.NKr.SARS-CoV-2.Spike cells. These cells were stained for viability (AquaVivid; Thermo Fisher Scientific) and cellular dye (cell proliferation dye eFluor670; Thermo Fisher Scientific) to be used as target cells. Overnight rested PBMCs from healthy donors were stained with another cellular marker (cell proliferation dye eFluor450; Thermo Fisher Scientific) and used as effector cells. Stained target and effector cells were mixed at a ratio of 1:10 in 96-well V-bottom plates. Plasma (1/500 dilution) or monoclonal antibodies CR3022 and CV3-13 (1 µg/mL) were added to the appropriate wells.^4^ The plates were subsequently centrifuged for 1 min at 300xg, and incubated at 37°C, 5% CO2 for 5 hours before being fixed in a 2% PBS-formaldehyde solution. ADCC activity (ADCC%) was calculated using the formula: [(% of GFP+ cells in Targets plus Effectors)-(% of GFP+ cells in Targets plus Effectors plus plasma/antibody)]/(% of GFP+ cells in Targets) x 100 by gating on transduced live target cells. Plasma from previous experiments and two monoclonal antibodies (CR3022, CV3-13) were used as quality controls and to standardize the ADCC ratio between experiments. All samples were acquired on an LSRII cytometer (BD Biosciences) and data analysis was performed using FlowJo v10.7.1 (Tree Star).

#### Plaque reduction neutralization test (PRNT) assay

The PRNT assay at NML was performed as previously published.^5^ SARS-CoV-2 (Canada/ON_ON-VIDO-01-2/2020, EPI_ISL_42517) stocks were titrated for use in the PRNT. Serological specimens were diluted 2-fold from 1:20 to 1:640 in DMEM supplemented with 2% FBS and incubated with 50 PFU of SARS-CoV-2 at 37°C and 5% CO2 for 1 hour. The sera-virus mixtures were added to 12-well plates containing Vero E6 cells at 100% confluence, followed by incubation at 37°C and 5% CO2 for 1 hour. After adsorption, a liquid overlay composed of 1.5% carboxymethylcellulose diluted in MEM, supplemented with 4% FBS, L-glutamine, nonessential amino acids, and sodium bicarbonate, was added to each well; the plates were incubated at 37°C and 5% CO2 for 72 hours. The liquid overlay was removed, and the cells were fixed with 10% neutral-buffered formalin for 1 hour at room temperature. The monolayers were stained with 0.5% crystal violet for 10 minutes and washed with 20% ethanol. Plaques were enumerated and compared with controls. The highest serum dilution resulting in 50% and 90% reduction in plaques compared with controls were defined as the PRNT50 and PRNT90 endpoint titers, respectively.

## Supplementary tables

**eTable 1:** Reasons for exclusion from the intention to treat and per protocol populations.

|  | Convalescent Plasma<br>(n, %) | Standard of Care<br>(n, %) |
| --- | --- | --- |
| **All randomized patients** | **627** | **313** |
| Consent withdrawal | 2(0.3) | 0(0.0) |
| Lost to follow-up (data on primary outcome unavailable; these patients were discharged alive before day 30, and could not be contacted on day 30) | 11(1.8) | 6(1.9) |
| **Intention to treat population** | **614** | **307** |
| *Ineligible for the trial* |  |  |
| Not receiving supplemental oxygen at the time of randomization (on oxygen at screening) | 4(0.6) | 4(1.3) |
| Any symptoms onset >12 days prior to randomization<sup>a</sup> | 1(0.2) | 0(0.0) |
| *Protocol deviation* |  |  |
| CCP arm received no CCP |  |  |
| Received regular plasma | 1(0.2) |  |
| Investigator decision | 9(1.4) |  |
| Patient refused | 7(1.1) |  |
| Total volume transfused < 400 ml | 11(1.8) |  |
| CCP transfusion ended > 24 hour post randomization | 33(5.3) |  |
| **Per protocol population** | **548(87.4)** | **303(96.8)** |

<sup>a</sup> For protocol version 5.0 or earlier (post-randomization, patient revised date of onset of symptoms)

**eTable 2:** Baseline characteristics of the intention to treat population. Excluding the 19 patients lost to follow-up at day 30 or withdrawing consent before day 30. Categorical data presented as number (percentage) and continuous variables as mean ± standard deviation and median (interquartile range).

| Characteristic | Convalescent Plasma<br>n=614 | Standard of Care<br>n=307 | Overall<br>n=921 |
| --- | --- | --- | --- |
| **Age - yr** | 67.8±16.0; 69 (58,80) | 67.3±14.8; 68 (58,78) | 67.6±15.6; 69 (58,79) |
| $\geq 60$ years | 433(70.5) | 216(70.4) | 649(70.5) |
| **Sex** |  |  |  |
| Male | 362(59.0) | 183(59.6) | 545(59.2) |
| **Pregnant at randomization** | 4(0.7) |  | 4(0.4) |
| **Ethnicity** |  |  |  |
| White | 101(16.4) | 44(14.3) | 145(15.7) |
| Asian | 300(48.9) | 151(49.2) | 451(49.0) |
| Hispanic or Latino | 25(4.1) | 11(3.6) | 36(3.9) |
| Black | 33(5.4) | 9(2.9) | 42(4.6) |
| Other | 37(6.0) | 28(9.1) | 65(7.1) |
| Unknown | 118(19.2) | 64(20.8) | 182(19.8) |
| **ABO blood group** |  |  |  |
| O | 265(43.2) | 112(36.5) | 377(40.9) |
| A | 231(37.6) | 119(38.8) | 350(38.0) |
| B | 87(14.2) | 54(17.6) | 141(15.3) |
| AB | 31(5.0) | 22(7.2) | 53(5.8) |
| **BMI – kg/m<sup>2</sup>** | 30.1±7.5; 29 (25,33) | 30.0±7.5; 29 (25,33) | 30.1±7.5; 29 (25,33) |
| BMI <30 | 248(40.4) | 121(39.4) | 369(40.1) |
| BMI $\geq 30$ | 198(32.2) | 100(32.6) | 298(32.4) |
| Unknown | 168(27.4) | 86(28.0) | 254(27.6) |
| **Smoking status** |  |  |  |
| Never Smoking | 324(52.8) | 159(51.8) | 483(52.4) |
| Former Smoker | 188(30.6) | 104(33.9) | 292(31.7) |
| Current Smoker | 19(3.1) | 6(2.0) | 25(2.7) |
| Unknown | 83(13.5) | 38(12.4) | 121(13.1) |
| **Smoking history** |  |  |  |
| <15 years or non-smoker | 355(57.8) | 172(56.0) | 527(57.2) |
| ≥15 years | 84(13.7) | 53(17.3) | 137(14.9) |
| Smokers with unknown history | 92(15.0) | 44(14.3) | 136(14.8) |
| Unknown | 83(13.5) | 38(12.4) | 121(13.1) |
| **Presence of comorbidity** |  |  |  |
| Diabetes | 214(34.9) | 104(33.9) | 318(34.5) |
| Cardiac disease | 379(61.7) | 193(62.9) | 572(62.1) |
| Baseline respiratory diseases | 145(23.6) | 79(25.7) | 224(24.3) |
| **Abnormal CT chest or chest x-ray result before randomization** | 553(90.1) | 261(85.0) | 814(88.4) |
| **Medication for other research study at baseline** |  |  |  |
| No | 562(91.5) | 267(87.0) | 829(90.0) |
| Yes | 52(8.5) | 40(13.0) | 92(10.0) |
| **Medication for COVID-19 at baseline** |  |  |  |
| Azithromycin | 277(45.1) | 134(43.6) | 411(44.6) |
| Other antibiotics | 400(65.1) | 182(59.3) | 582(63.2) |
| Steroid/Immunomodulator | 489(79.6) | 244(79.5) | 733(79.6) |
| Antiviral medications | 161(26.2) | 78(25.4) | 239(26.0) |
| Anticoagulants | 345(56.2) | 175(57.0) | 520(56.5) |
| Other COVID-19 Medications | 76(12.4) | 36(11.7) | 112(12.2) |
| **Medication not for COVID-19 at baseline** |  |  |  |
| ACE inhibitor | 84(13.7) | 62(20.2) | 146(15.9) |
| ACE receptor blocker(ARB) | 76(12.4) | 46(15.0) | 122(13.2) |
| Non-steroidal anti-inflammatory drugs(NSAIDs) | 77(12.5) | 51(16.6) | 128(13.9) |
| Colchicine | 5(0.8) | 2(0.7) | 7(0.8) |
| Systemic corticosteroids | 60(9.8) | 35(11.4) | 95(10.3) |
| Inhaled corticosteroids | 82(13.4) | 42(13.7) | 124(13.5) |
| Immunomodulatory agents | 22(3.6) | 17(5.5) | 39(4.2) |
| Anticoagulants | 132(21.5) | 63(20.5) | 195(21.2) |
| Oxygen status at baseline (FiO2) | 49.7±25.3; 40 (30,65) | 49.1±25.3; 40 (30,60) | 49.5±25.3; 40 (30,64) |
| Time from first Covid-19 symptom to randomization (days) | 8.0±3.8; 8 (5,10) | 7.8±3.4; 8 (5,10) | 7.9±3.7; 8 (5,10) |
| Time from Covid-19 diagnosis<sup>a</sup> to randomization (days) | 5.0±3.6; 4 (2,8) | 5.0±4.4; 4 (2,7) | 5.0±3.9; 4 (2,7) |
| Enrolled in other clinical trials | 166(27.0) | 96(31.3) | 262(28.4) |

<sup>a</sup>Day of positive Covid-19 test.

**eTable 3:**
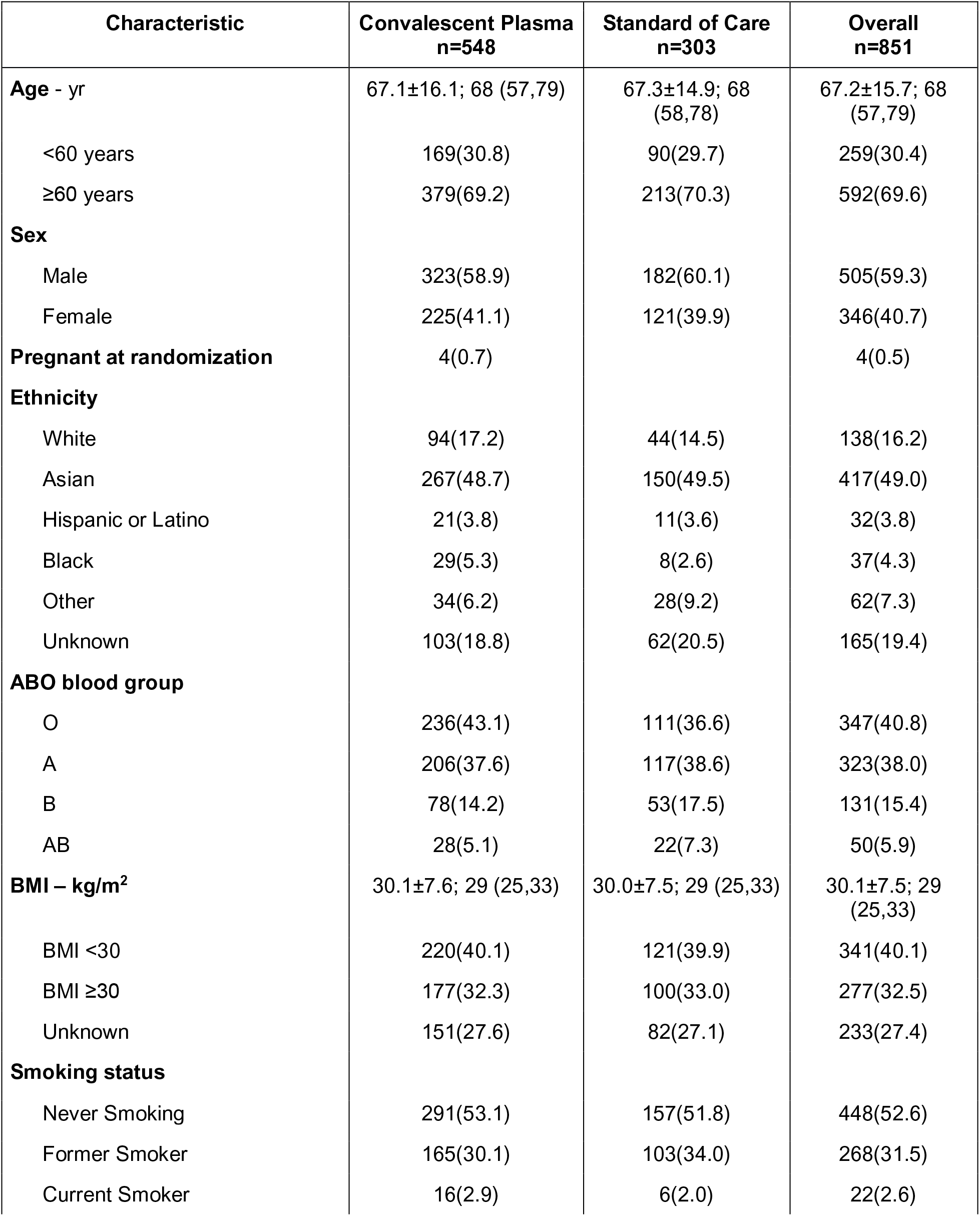
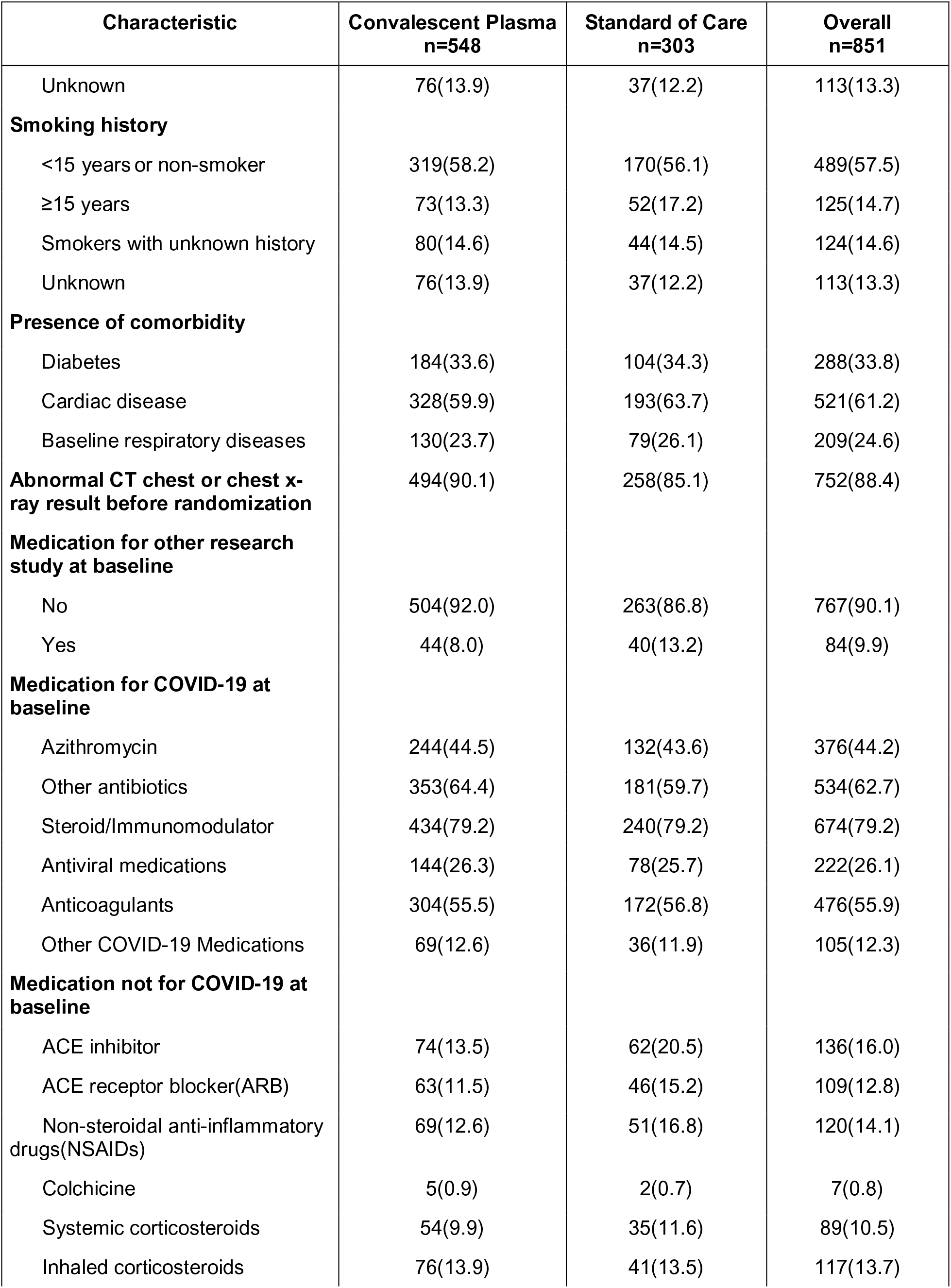
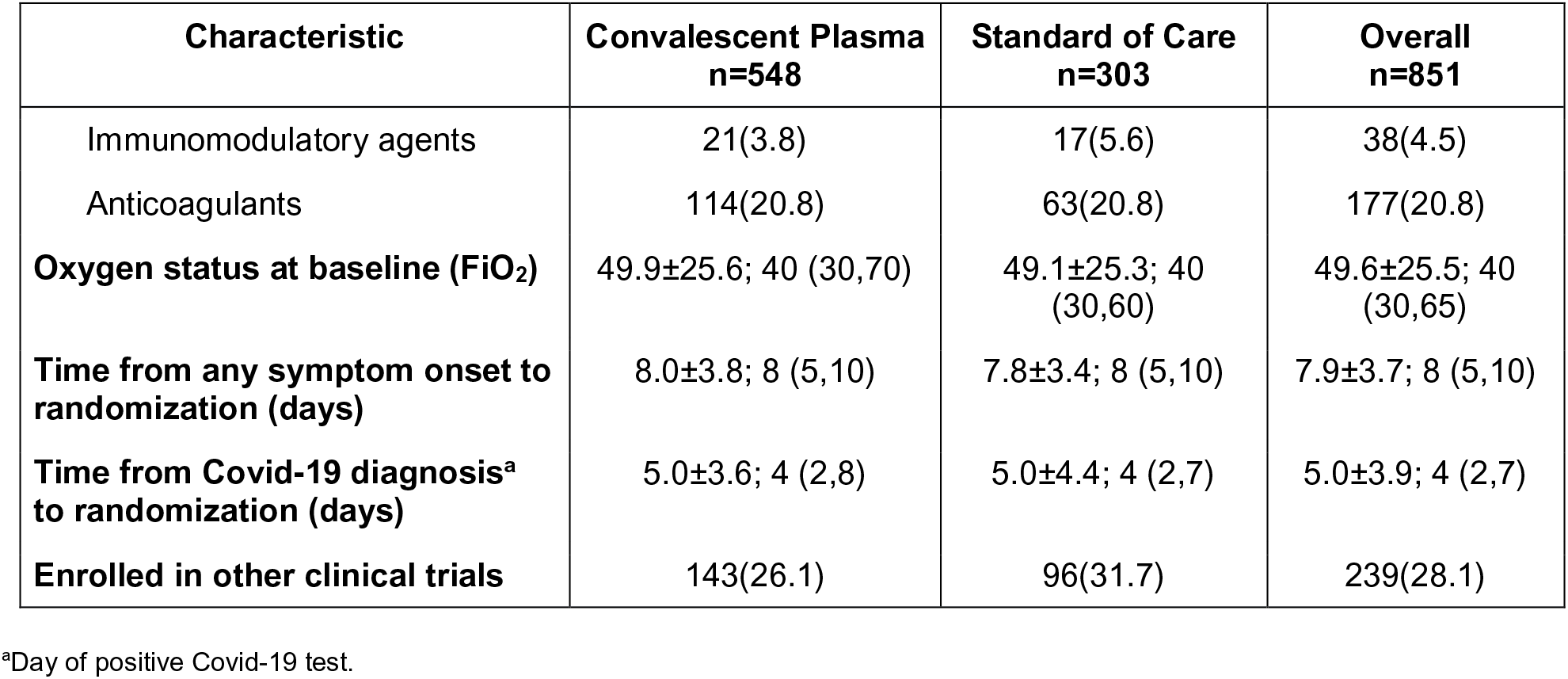
Baseline characteristics of the per protocol population. Categorical data presented as number (percentage) and continuous variables as mean ± standard deviation and median (interquartile range).

**eTable 4:** Primary and secondary end points for the per protocol population.

| Per protocol analysis | Population | CCP<br>n=548 | SOC<br>n=303 | Treatment effect<sup>a</sup> |
| --- | --- | --- | --- | --- |
| **Intubation or death at day30** | **851<sup>b</sup>** | **167(30.5)** | **85(28.1)** | **RR=1.09 (0.87,1.35),**<br>**p=0.4612** |
| Time to Intubation or in-hospital death<br>(days) by day30 | 851 |  |  | HR=1.062 (0.819,1.376),<br>p=0.6516 |
| Number of ventilation-free days<br>by day30 | 851 | 23.9±10.2 | 24.0±10.5 | MD=-0.1 (-1.5, 1.3),<br>p=0.8520 |
| Any death by day30 | 851 | 114(20.8) | 62(20.5) | RR=1.02 (0.77,1.34),<br>p=0.9065 |
| Length of stay in ICU by day 30 | 851 | 4.3±8.0 | 3.7±7.1 | MD=0.6 (-0.4, 1.7),<br>p=0.2485 |
| Need for Renal replacement<br>therapy by<br>day30<sup>c</sup> | 842 | 7(1.3) | 6(2.0) | RR=0.64 (0.22,1.89),<br>p=0.4225 |
| Serious Adverse event by day30 | 851 | 172(31.4) | 80(26.4) | RR=1.19 (0.95,1.49),<br>p=0.1321 |
| **Per protocol analysis** |  | **n=557** | **n=309** | **Treatment effect** |
| In-hospital death by day 90 | 866<sup>d</sup> | 127(22.8) | 68(22.0) | RR=1.04 (0.80,1.34),<br>p=0.7890 |
| Time to in-hospital death by day<br>90 | 866 |  |  | HR=0.946 (0.704,1.270),<br>p=0.946 |
| Length of stay in hospital by day<br>90 | 866 |  |  | HR=0.930 (0.810,1.067),<br>p=0.2999 |

<sup>a</sup> Relative Risk (RR, 95% confidence interval), Hazard Ratio (HR, 95% confidence interval) and Mean Difference (MD, with 95% CI based on robust bootstrap standard errors)
<sup>b</sup>See Figure 1 for exclusions to the per protocol population
<sup>c</sup>Excluding 9 patients on chronic kidney replacement therapy at baseline
<sup>d</sup>Including 15 additional patients above the per-protocol, n=851, who were lost to follow-up after discharge from hospital

**eTable 5:** Antibody test results overall and by participating blood supplier.

|  | CCP units<br>n=1192 | Concentration<br>Median (Q1,Q3) (min, max) |
| --- | --- | --- |
| **ADCC ratio<sup>a</sup>** |  |  |
| All sites | 1186 (99.5) | 1.359 (0.603, 3.051) (0.127,15.481) |
| Supplier 1 | 553 (99.1) | 2.284 (0.788, 4.375) (0.127,15.481) |
| Supplier 2 | 454 (100.0) | 1.122 (0.571, 2.437) (0.127,10.321) |
| Supplier 3 | 174 (100.0) | 0.583 (0.351, 1.091) (0.127,3.568) |
| Supplier 4 | 5 (83.3) | 0.844 (0.599, 1.506) (0.196,1.795) |
| **Anti-RBD ELISA<sup>b</sup>** |  |  |
| All sites | 1186 (99.5) | 1.410 (0.959, 1.836) (0.215, 2.943) |
| Supplier 1 | 553 (99.1) | 1.638 (1.342, 2.029) (0.215, 2.829) |
| Supplier 2 | 454 (100.0) | 1.102 (0.668, 1.678) (0.304, 2.943) |
| Supplier 3 | 174 (100.0) | 1.131 (0.810, 1.580) (0.232, 2.423) |
| Supplier 4 | 5 (83.3) | 1.321 (1.171, 1.718) (0.758, 2.011) |
| **Anti-S IgG<sup>c</sup>** |  |  |
| All sites | 1186 (99.5) | 2853 (1372, 4967) (117, 30402) |
| Supplier 1 | 553 (99.1) | 3530 (2116, 5547) (117, 15506) |
| Supplier 2 | 454 (100.0) | 2308 (1248, 4967) (261, 30402) |
| Supplier 3 | 174 (100.0) | 1333 (578, 2469) (193, 9271) |
| Supplier 4 | 5 (83.3) | 2477 (1974,2777) (700,10428) |
| **PRNT-50<sup>d,e</sup>** |  |  |
| All sites | 1191(99.9) | 160 (80,320) (10,1280) |
| Supplier 1 | 558 (100.0) | 160 (160, 640) (40, 1280) |
| Supplier 2 | 454(100.0) | 80 (20,160) (10, 640) |
| Supplier 3 | 174(100.0) | 80 (30,160) (10, 640) |
| Supplier 4 | 5 (83.3) | 160 (160, 160) (80, 160) |

<sup>a</sup>Mean of ADCC ratio (1-7 measures). ADCC % with limit 11.3% (equivalent to ratio 0.127); ADCC ratio=ADCC %/(100 -ADCC%)
<sup>b</sup>Mean of anti-RBD ELISA conc. (1-8 measures)
<sup>c</sup>Mean of anti-S IgG conc. (1-7 measures)
<sup>d</sup>Median of PRNT-50 (1-6 measures): PRNT-50 titer=1: n then PRNT=n, PRNT-50 titer≥1:1280 then PRNT=1280, PRNT-50 titer≥1:640 then PRNT=640, PRNT-50 titer=negative then PRNT=10
<sup>e</sup>Includes interpolations (NYBC n=6 and CBS n=1)

**eTable 6:** Adverse events at day 30 for the intention to treat population. Categorical data presented as number (percentage) and continuous variables as mean ± standard deviation and median (interquartile range).

| Intention to treat analysis | Convalescent Plasma<br>n=614 | Standard of Care<br>n=307 | Overall<br>n=921 |
| --- | --- | --- | --- |
| **Patients with adverse events** |  |  |  |
| Grade $\geq 3$ (Severe) | 260(42.3) | 109(35.5) | 369(40.1) |
| Grade $\geq 4$ (Life-Threatening) | 188(30.6) | 74(24.1) | 262(28.4) |
| Grade 5 (Fatal) | 141 (23.0) | 63 (20.5) | 204 (22.1) |
| **Number of adverse events per patient** |  |  |  |
| Grade $\geq 3$ (Severe) | 0.9±1.7; 0 (0,1) | 0.6±1.2; 0 (0,1) | 0.8±1.5; 0 (0,1) |
| Grade $\geq 4$ (Life-Threatening) | 0.4±0.7; 0 (0,1) | 0.3±0.7; 0 (0,0) | 0.4±0.7; 0 (0,1) |
| Grade 5 (Fatal) | 0.2±0.4; 0 (0,0) | 0.2±0.4; 0 (0,0) | 0.2±0.4; 0 (0,0) |
| **Serious adverse event (grade $\geq 3$)** | 205 (33.4) | 81 (26.4) | 286 (31.1) |
| **Number of SAEs per patient** | 0.4±0.8; 0 (0,1) | 0.3±0.7; 0 (0,1) | 0.4±0.7; 0 (0,1) |
| **Grade 3 non CCP related adverse event** |  |  |  |
| Hypoxia | 23 (3.7) | 7 (2.3) | 30 (3.3) |
| Lymphocyte count decreased | 17 (2.8) | 8 (2.6) | 25 (2.7) |
| Infections and infestations - Other, specify | 15 (2.4) | 8 (2.6) | 23 (2.5) |
| Anemia | 15 (2.4) | 3 (1.0) | 18 (2.0) |
| Hyperglycemia | 13 (2.1) | 5 (1.6) | 18 (2.0) |
| Hypertension | 11 (1.8) | 2 (0.7) | 13 (1.4) |
| Pneumonitis | 10 (1.6) | 1 (0.3) | 11 (1.2) |
| Hypermagnesemia | 9 (1.5) | 1 (0.3) | 10 (1.1) |
| **Grade 4 non CCP related adverse event** |  |  |  |
| Hypoxia | 22 (3.6) | 6 (2.0) | 28 (3.0) |
| Acute kidney injury | 12 (2.0) | 7 (2.3) | 19 (2.1) |
| Sepsis | 8 (1.3) | 4 (1.3) | 12 (1.3) |
| **Grade 5 non CCP related adverse event** |  |  |  |
| Respiratory failure | 63 (10.3) | 32 (10.4) | 95 (10.3) |
| Hypoxia | 30 (4.9) | 15 (4.9) | 45 (4.9) |
| Death<sup>a</sup> | 14 (2.3) | 4 (1.3) | 18 (2.0) |
| **Grade $\geq 3$ non CCP related adverse event** |  |  |  |
| Respiratory failure | 76 (12.4) | 35 (11.4) | 111 (12.1) |
| Hypoxia | 75 (12.2) | 28 (9.1) | 103 (11.2) |
| Lymphocyte count decreased | 17 (2.8) | 8 (2.6) | 25 (2.7) |
| Infections and infestations - Other | 16 (2.6) | 8 (2.6) | 24 (2.6) |
| Acute kidney injury | 13 (2.1) | 10 (3.3) | 23 (2.5) |
| Sepsis | 14 (2.3) | 7 (2.3) | 21 (2.3) |
| Anemia | 16 (2.6) | 3 (1.0) | 19 (2.1) |
| Death<sup>a</sup> | 14 (2.3) | 4 (1.3) | 18 (2.0) |
| Hyperglycemia | 13 (2.1) | 5 (1.6) | 18 (2.0) |
| Adult respiratory distress syndrome | 10 (1.6) | 3 (1.0) | 13 (1.4) |
| Dyspnea | 9 (1.5) | 4 (1.3) | 13 (1.4) |
| Hypertension | 11 (1.8) | 2 (0.7) | 13 (1.4) |
| Hypotension | 10 (1.6) | 3 (1.0) | 13 (1.4) |
| Respiratory, thoracic, mediastinal disorders | 10 (1.6) | 3 (1.0) | 13 (1.4) |
| Lung infection | 9 (1.5) | 4 (1.3) | 13 (1.4) |
| Thromboembolic event | 7 (1.1) | 5 (1.6) | 12 (1.3) |
| Pneumonitis | 10 (1.6) | 1 (0.3) | 11 (1.2) |
| Hypermagnesemia | 9 (1.5) | 1 (0.3) | 10 (1.1) |

<sup>a</sup>Adverse event classified by the investigator as CTCAE term "death (not otherwise specified)".

**eTable 7:** Adverse events at day 30 for the per protocol population. Categorical data presented as number (percentage) and continuous variables as mean ± standard deviation and median (interquartile range).

| Per protocol analysis | Convalescent Plasma<br>n=548 | Standard of Care<br>n=303 | Overall<br>n=851 |
| --- | --- | --- | --- |
| **Patients with adverse events** |  |  |  |
| Grade $\geq 3$ (Severe) | 218(39.8) | 108(35.6) | 326(38.3) |
| Grade $\geq 4$ (Life-Threatening) | 156(28.5) | 73(24.1) | 229(26.9) |
| Grade $\geq 5$ (Fatal) | 114 (20.8) | 62 (20.5) | 176 (20.7) |
| **Number of adverse events per patient** |  |  |  |
| Grade $\geq 3$ (Severe) | 0.8±1.7; 0 (0,1) | 0.6±1.2; 0 (0,1) | 0.7±1.5; 0 (0,1) |
| Grade $\geq 4$ (Life-Threatening) | 0.4±0.7; 0 (0,1) | 0.3±0.7; 0 (0,0) | 0.3±0.7; 0 (0,1) |
| Grade 5 (Fatal) | 0.2±0.4; 0 (0, 0) | 0.2±0.4; 0 (0, 0) | 0.2±0.4; 0 (0, 0) |
| **Serious adverse event** | 172 (31.4) | 80 (26.4) | 252 (29.6) |
| **Number of SAEs per patient** | 0.4±0.7; 0 (0, 1) | 0.3±0.7; 0 (0, 1) | 0.4±0.7; 0 (0, 1) |
| **Grade 3 non CCP related adverse event** |  |  |  |
| Hypoxia | 20 (3.6) | 7 (2.3) | 27 (3.2) |
| Infections and infestations - Other, specify | 14 (2.6) | 8 (2.6) | 22 (2.6) |
| Lymphocyte count decreased | 14 (2.6) | 8 (2.6) | 22 (2.6) |
| Anemia | 14 (2.6) | 3 (1.0) | 17 (2.0) |
| Hyperglycemia | 11 (2.0) | 5 (1.7) | 16 (1.9) |
| Hypertension | 10 (1.8) | 2 (0.7) | 12 (1.4) |
| Pneumonitis | 10 (1.8) | 1 (0.3) | 11 (1.3) |
| Hypermagnesemia | 8 (1.5) | 1 (0.3) | 9 (1.1) |
| Hypophosphatemia | 8 (1.5) | 1 (0.3) | 9 (1.1) |
| Urinary tract infection | 7 (1.3) | 2 (0.7) | 9 (1.1) |
| **Grade 4 non CCP related adverse event** |  |  |  |
| Hypoxia | 20 (3.6) | 6 (2.0) | 26 (3.1) |
| Acute kidney injury | 11 (2.0) | 7 (2.3) | 18 (2.1) |
| Sepsis | 8 (1.5) | 4 (1.3) | 12 (1.4) |

| Per protocol analysis | Convalescent<br>Plasma<br>n=548 | Standard of Care<br>n=303 | Overall<br>n=851 |
| --- | --- | --- | --- |
| **Grade 5 non CCP related adverse event** |  |  |  |
| Respiratory failure | 53 (9.7) | 31 (10.2) | 84 (9.9) |
| Hypoxia | 25 (4.6) | 15 (5.0) | 40 (4.7) |
| Death<sup>a</sup> | 11 (2.0) | 4 (1.3) | 15 (1.8) |
| **Grade 3/4/5 non CCP related adverse event** |  |  |  |
| Respiratory failure | 64 (11.7) | 34 (11.2) | 98 (11.5) |
| Hypoxia | 65 (11.9) | 28 (9.2) | 93 (10.9) |
| Infections and infestations - Other, specify | 14 (2.6) | 8 (2.6) | 22 (2.6) |
| Lymphocyte count decreased | 14 (2.6) | 8 (2.6) | 22 (2.6) |
| Acute kidney injury | 12 (2.2) | 10 (3.3) | 22 (2.6) |
| Sepsis | 13 (2.4) | 7 (2.3) | 20 (2.4) |
| Anemia | 15 (2.7) | 3 (1.0) | 18 (2.1) |
| Hyperglycemia | 11 (2.0) | 5 (1.7) | 16 (1.9) |
| Death<sup>a</sup> | 11 (2.0) | 4 (1.3) | 15 (1.8) |
| Dyspnea | 8 (1.5) | 4 (1.3) | 12 (1.4) |
| Hypertension | 10 (1.8) | 2 (0.7) | 12 (1.4) |
| Respiratory, thoracic and mediastinal disorders | 9 (1.6) | 3 (1.0) | 12 (1.4) |
| Lung infection | 8 (1.5) | 4 (1.3) | 12 (1.4) |
| Pneumonitis | 10 (1.8) | 1 (0.3) | 11 (1.3) |
| Thromboembolic event | 6 (1.1) | 5 (1.7) | 11 (1.3) |
| Adult respiratory distress syndrome | 7 (1.3) | 3 (1.0) | 10 (1.2) |
| Hypotension | 7 (1.3) | 3 (1.0) | 10 (1.2) |
| Hypermagnesemia | 8 (1.5) | 1 (0.3) | 9 (1.1) |
| Hypophosphatemia | 8 (1.5) | 1 (0.3) | 9 (1.1) |
| Urinary tract infection | 7 (1.3) | 2 (0.7) | 9 (1.1) |

<sup>a</sup>Adverse event classified by the investigator as CTCAE term "death (not otherwise specified)".

**eTable 8:** Convalescent plasma adverse events at day 30 for the intention to treat population.

| Intention to treat analysis | Convalescent Plasma<br>n=614 |
| --- | --- |
| CCP related adverse event, n |  |
| CTCAE Grade 1 | 10 (1.6) |
| CTCAE Grade 2 | 10 (1.6) |
| CTCAE Grade 3 (Severe) | 8 (1.3) |
| CTCAE Grade 4 (Life-Threatening) | 4 (0.7) |
| CTCAE Grade 5 (Fatal) |  |
| Number of CCP related AEs per patient* | 0.0±0.1; 0 (0, 0) |
| CTCAE Grade 1 | 0.0±0.1; 0 (0, 0) |
| CTCAE Grade 2 | 0.0±0.1; 0 (0, 0) |
| CTCAE Grade 3 (Severe) | 0.0±0.1; 0 (0, 0) |
| CTCAE Grade 4 (Life-Threatening) | 0.0±0.0; 0 (0, 0) |
| CTCAE Grade 5 (Fatal) |  |
| CCP related adverse event, n |  |
| ISBT Grade 1 | 18 (2.9) |
| ISBT Grade 2 | 10 (1.6) |
| ISBT Grade 3 | 4 (0.7) |
| ISBT Grade 4 |  |
| Number of CCP related AEs per patient<sup>a</sup> |  |
| ISBT Grade 1 | 0.0±0.2; 0 (0,0) |
| ISBT Grade 2 | 0.0±0.1; 0 (0,0) |
| ISBT Grade 3 | 0.0±0.1; 0 (0,0) |
| ISBT Grade 4 | 0.0±0.0; 0 (0,0) |
| Event name, n |  |
| TAD | 13 (2.1) |
| Minor Allergic Reaction | 9 (1.5) |
| TACO | 5 (0.8) |

| **Intention to treat analysis** | Convalescent<br>Plasma<br>n=614 |
| --- | --- |
| Febrile Non-Hemolytic Reaction | 4 (0.7) |
| Other (Hypertensive Reaction) | 2 (0.3) |
| Hypotensive Reaction | 1 (0.2) |
| Possible TRALI | 1 (0.2) |

<sup>a</sup>Mean/standard deviation); median (interquartile range)

**eTable 9:**
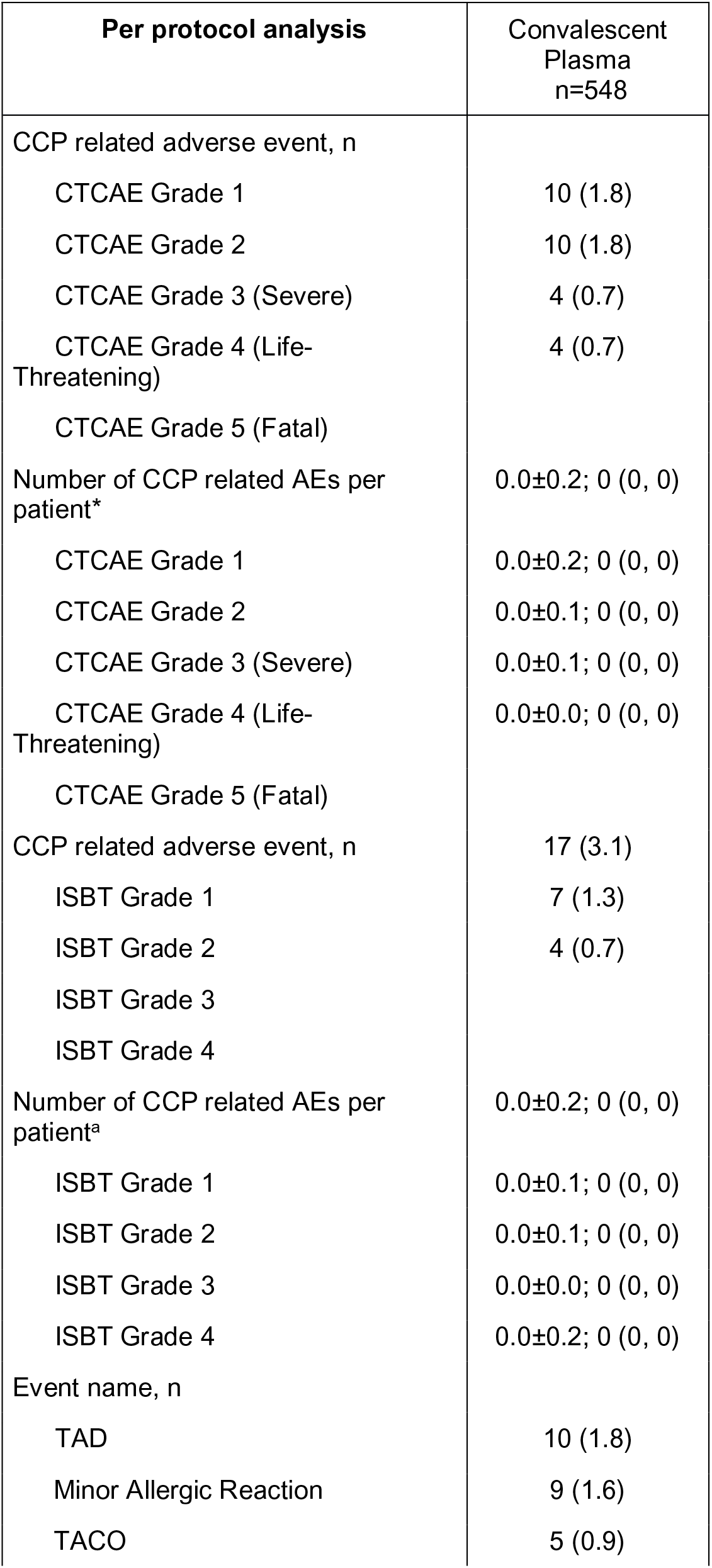
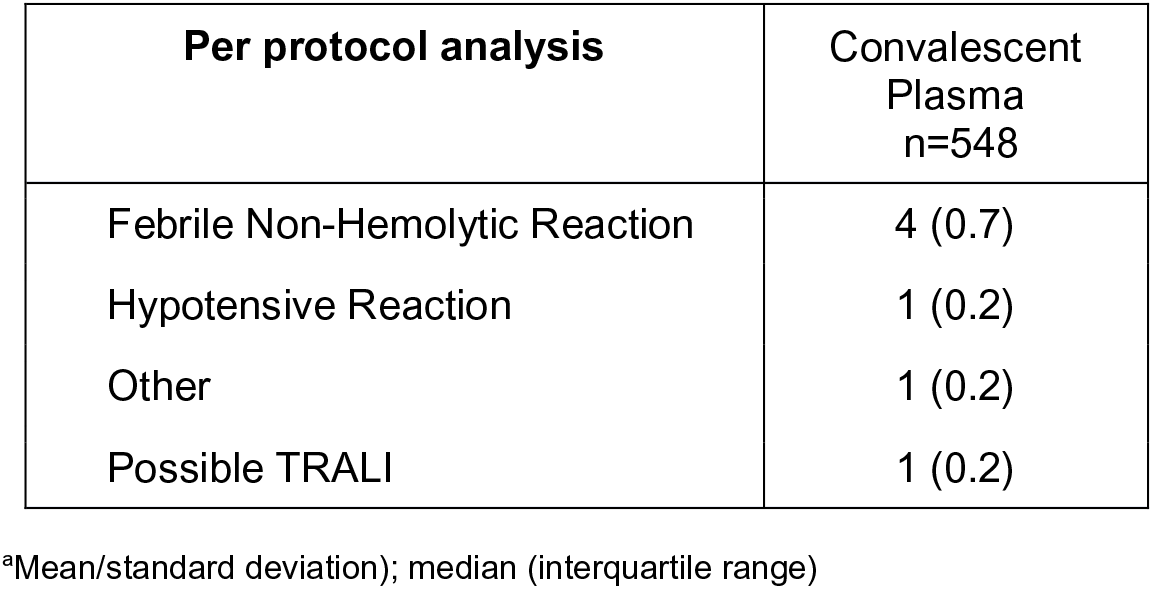
Convalescent plasma adverse events at day 30 for the per protocol population.

**eTable 10:**
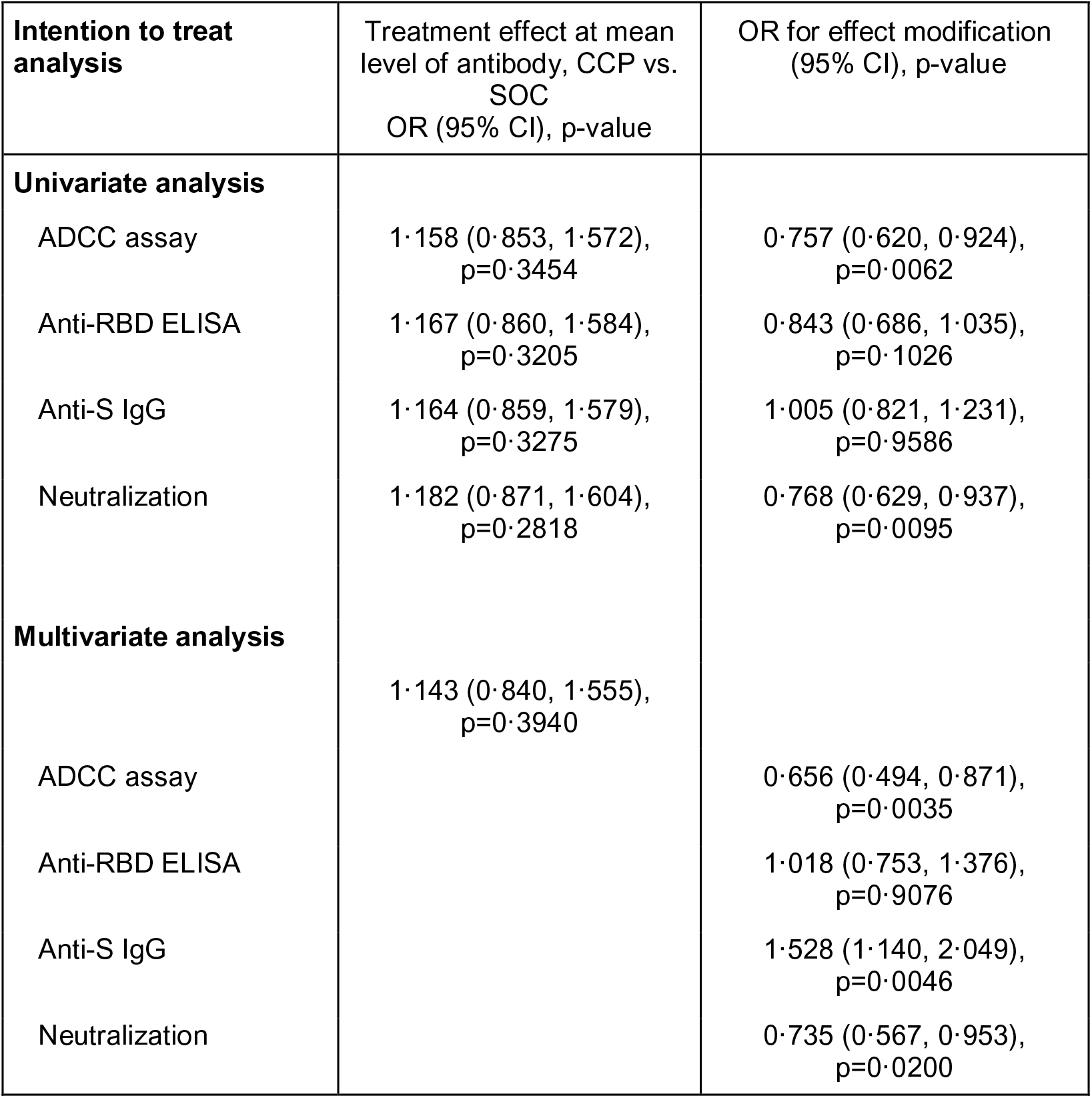
Summary of logistic regression models with standardized continuous log marker dose. The odds ratio reported in the third column reflects the effect modification corresponding to a 1 standard deviation unit increase in the log transformed and entered marker; in multivariate results markers other than the one labeled in the left column are set to mean log marker dose so the other covariates are zero. The convalescent plasma units transfused in the trial came from 530 distinct plasma donations. Five were missing all serologic marker values except for the original qualifying test and were excluded from analyses. Seven were missing only the neutralization titer value, which was interpolated from the other serologic markers, as described in the statistical analysis plan.

## Supplementary Figures

**eFigure 1:**
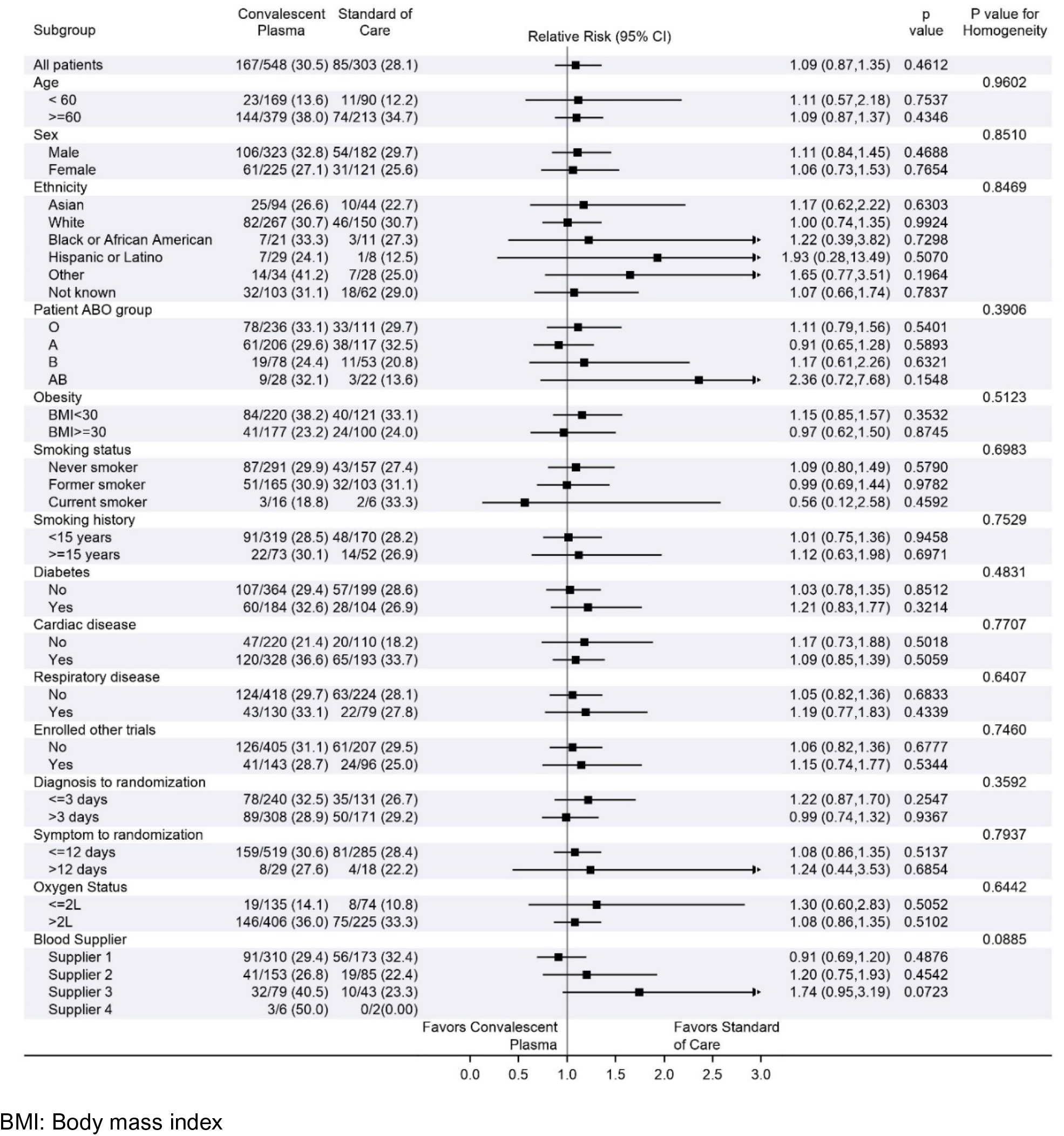
Subgroup analysis for the per-protocol population.

**eFigure 2:**
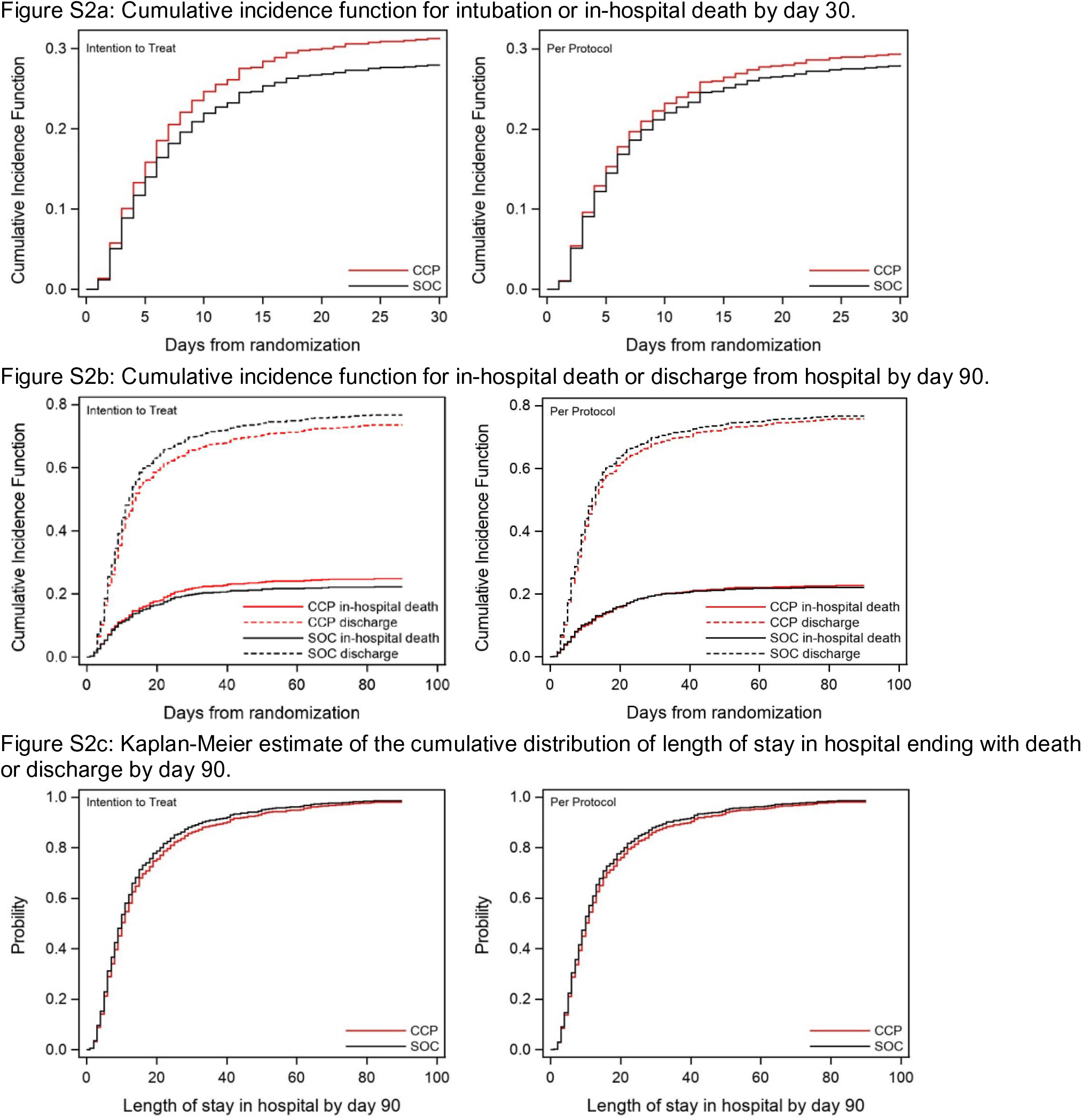
Cumulative incidence functions of intubation or in-hospital death by day 30 and in-hospital death by day 90, and Kaplan-Meier estimate of distribution of length of stay in hospital by day 90.

**eFigure 3:**
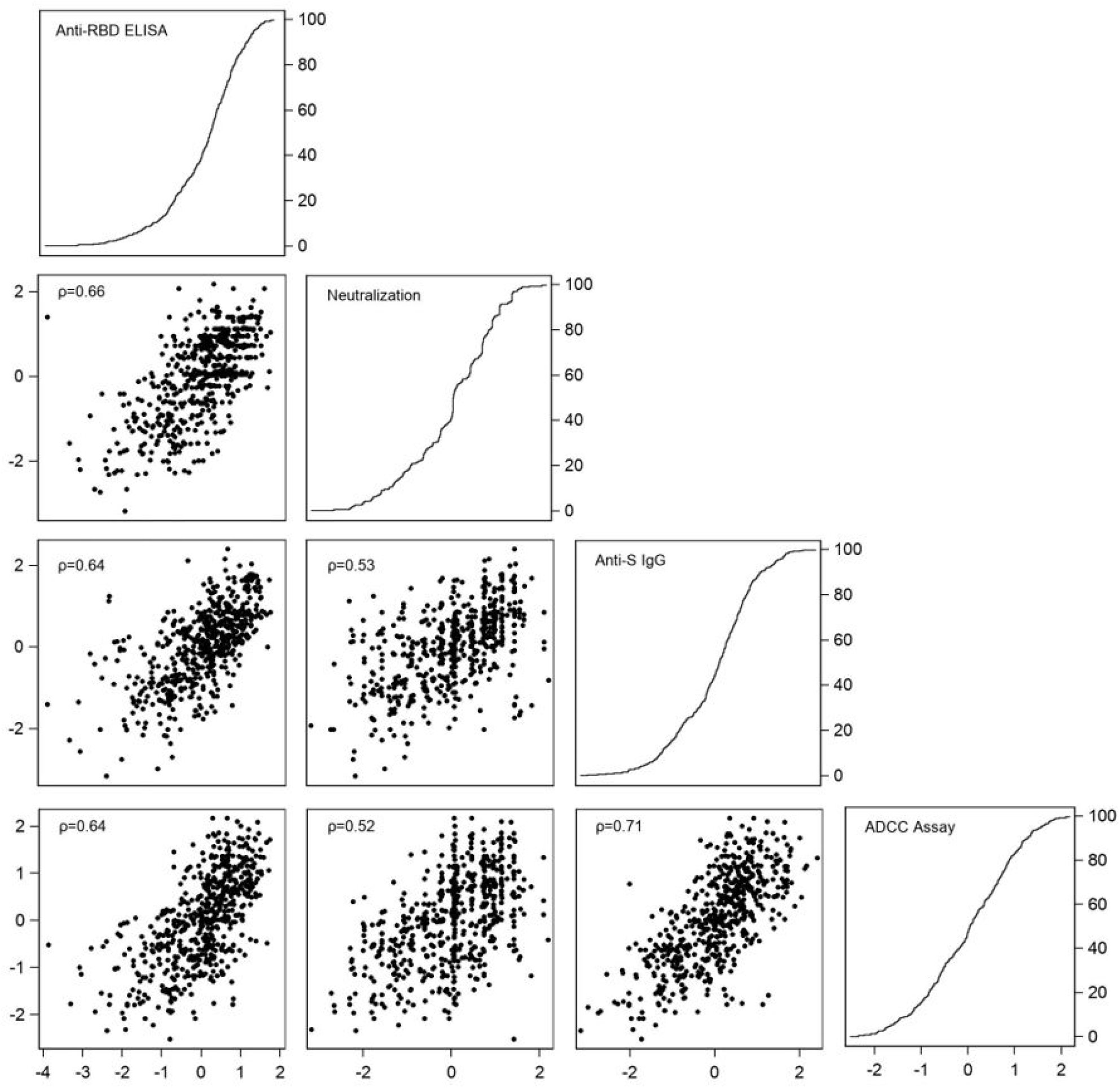
Pairwise scatter plots of plasma antibody markers and empirical distribution functions. Markers (log transformed and standardized) include Anti-RBD ELISA, plaque reduction neutralization test, Anti-S IgG and ADCC assay. (ρ: Pearson correlation coefficients of pair of antibody markers).

**eFigure 4:**
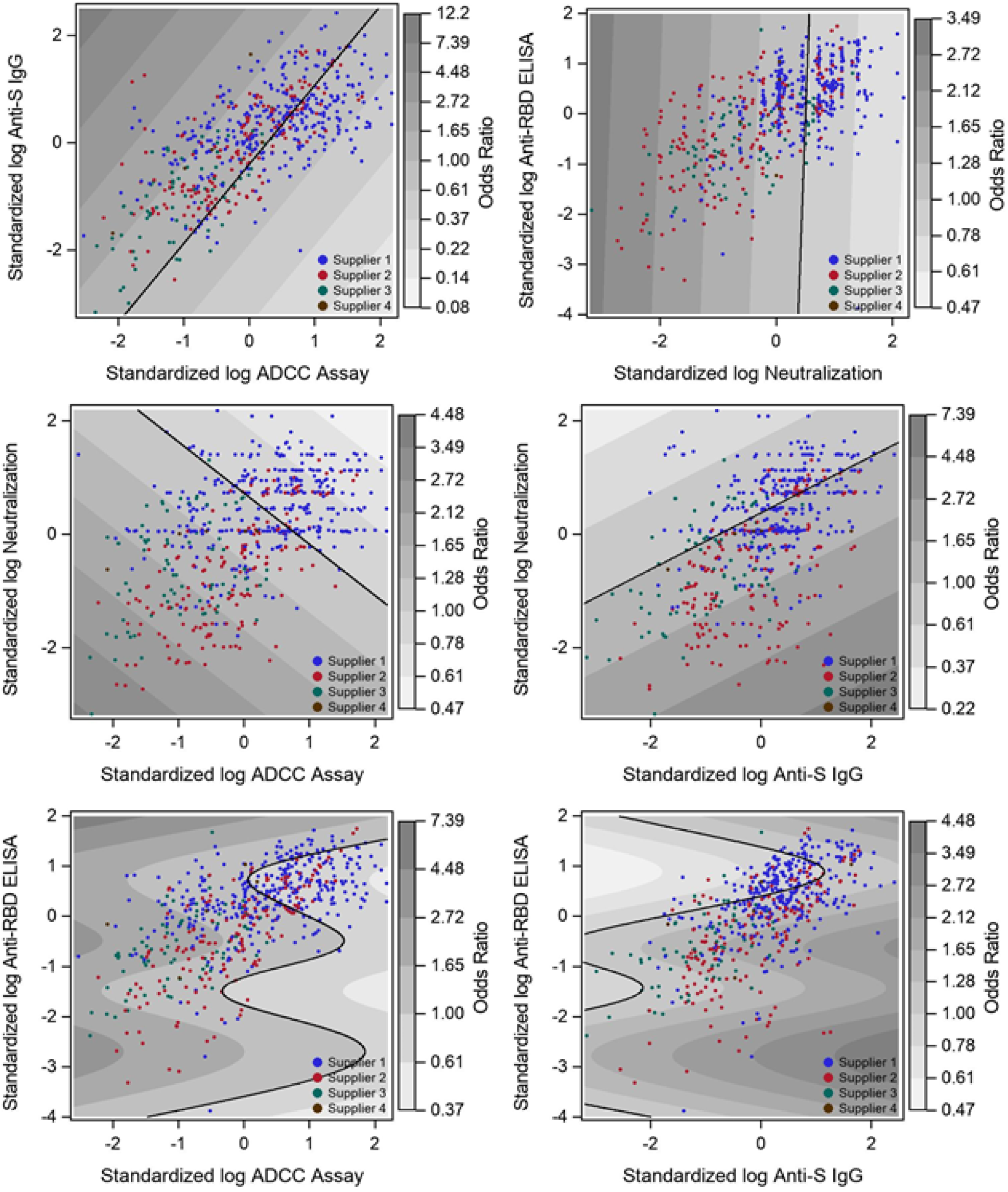
Exploration of the joint effect of antibody markers in convalescent plasma on the primary outcome. Contour plots of the odds ratio for the odds ratio for the composite event of intubation or death for individuals receiving blood as a function of the product of each possible pair of antibody markers. Over-layed data points indicate the value of the two antibody markers for each convalescent plasma transfusion in the study. The contours are obtained from a generalized additive logistic model for the primary outcome including blood supply center, treatment and the log transformed and standardized biomarkers using smoothing splines.

**eFigure 5:**
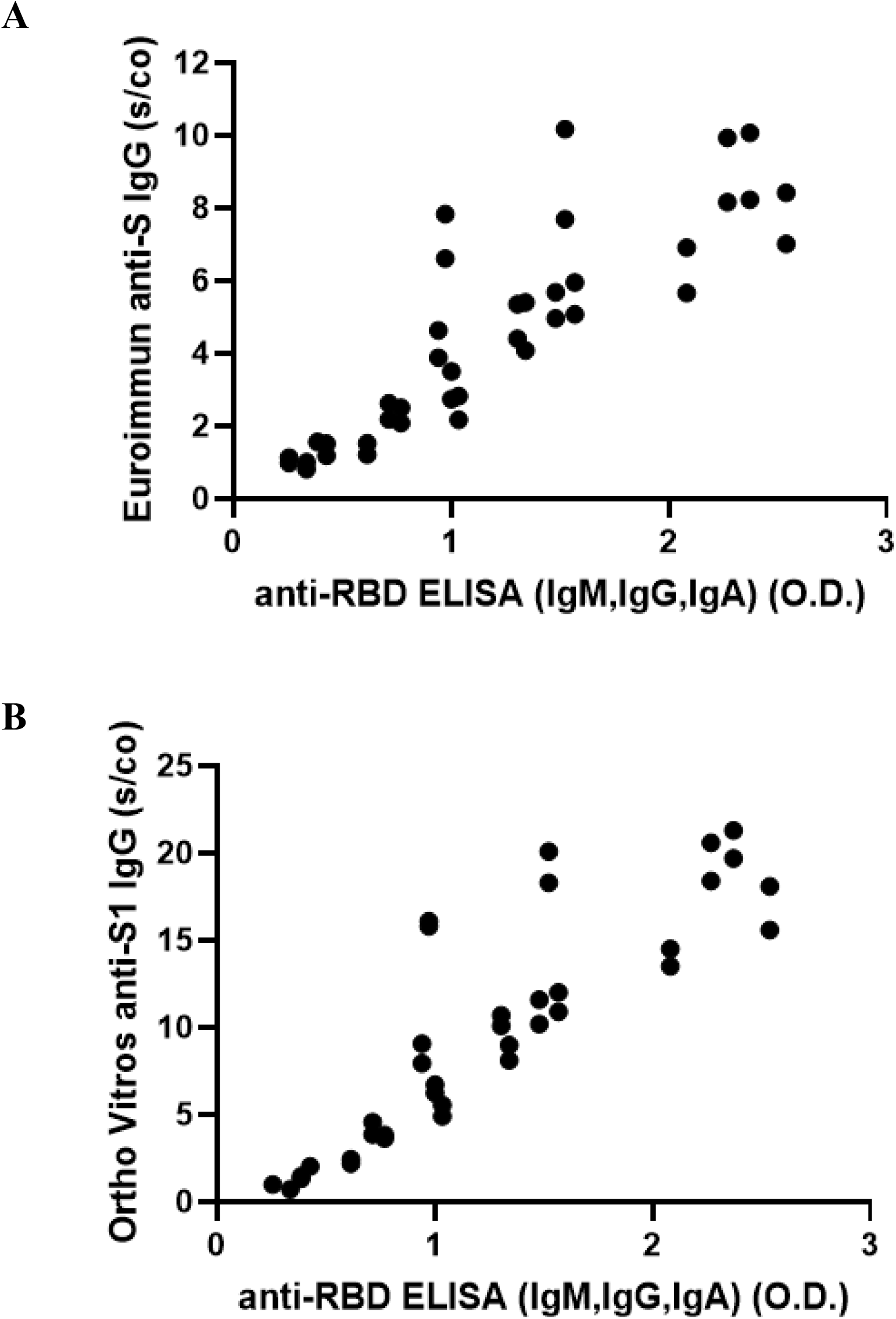
Comparison of in-house ELISA to commercial assays. Values from the Héma-Québec in-house ELISA measuring antibody (IgM, IgA, IgG) binding the receptor binding domain of SARS-CoV-2 Spike protein (used in the current study) are compared to results from A) Euroimmun and B) Ortho Vitros commercial assays measuring IgG binding to subunit 1 of the SARS-CoV-2 Spike protein, which contains the receptor binding domain and which were used to qualify convalescent plasma in previous clinical trials. Each sample was tested with the commercial assays twice.

## Notes

Grant Support: Canadian Institutes of Health Research – COVID-19 May 2020 Rapid Research Funding Opportunity – Operating Grant; Ontario COVID-19 Rapid Research Fund; Toronto COVID-19 Action Initiative 2020 (University of Toronto); University Health Network Emergent Access Innovation Fund; University Health Academic Health Science Centre Alternative Funding Plan (Sunnybrook Health Sciences Centre); Ministère de l’Économie et de l’Innovation; Fond de Recherche du Québec en Santé; Saskatchewan Ministry of Health; University of Alberta Hospital Foundation; Alberta Health Services COVID-19 Foundation Competition; Sunnybrook Health Sciences Centre Foundation; Fondations CHU Ste-Justine; The Ottawa Hospital Academic Medical Organization; The Ottawa Hospital Foundation COVID-19 Research Fund; Fondation du CHUM; Sinai Health System Foundation and McMaster University.

### Competing Interest Statement

PB reports payments to his institution from Canadian Institutes of Health Research, Ministere de l'economie et de l'innovation du Quebec, Fondation du CHU Sainte-Justine, Fondation du CHUM, and Fonds de Recherche du Quebec en Sante; JC reports research grants from Canadian Institutes of Health Research, University of Toronto, Sunnybrook Health Sciences Centre, and Canadian Blood Services; NMH reports research project funding unrelated to this study from Canadian Institutes of Health Research and Canadian Blood Services, honorarium for educational events unrelated to this study from CSL Behring Canada, and participation on a Data Safety Monitoring Board or Advisory Board for Canadian Blood Services (Scientific Advisory Committee); MPZ reports consulting fees from Canadian Blood Services in her role as a Medical Officer but receives no direct benefit from the study results; GBB reports a grant from Canadian Institutes of Health Research for Masters students; MMC reports consulting fees from Octapharma, participation in a data safety monitoring board or advisory board for Cerus and Haemonetics, and receipt of equipment, materials, drugs, medical writing, gifts or other services by her institution from Cerus; ND reports grants from AFP Innovation Fund Award Sunnybrook Health Sciences Centre, and University of Toronto; MJG reports payments to his institution from Regeneron, consulting fees from Regeneron and ReAlta Life Sciences, and participation on a Data Safety Monitoring Board or Advisory Board for Enzychem and Sobi; BSS reports employment by New York Blood Centre (but receiving no direct benefit from study participation); LS reports a leadership or fiduciary role in other board, society, committee or advocacy group, paid or unpaid (Member of the Ontario Bioethics Table for COVID-19); NS reports payment to her institution from the MSH Foundation for CONCOR; AF reports funding from Fondation du CHUM, Ministere de l'economie et de l'Innovation du Quebec, and Canadian Institutes of Health Research. All other authors declare no competing interests.

### Clinical Trial

NCT04348656

### Clinical Protocols

https://trialsjournal.biomedcentral.com/articles/10.1186/s13063-021-05235-3

### Author Declarations

The study was approved by Clinical Trials Ontario (Research Ethics Board of Record: Sunnybrook Health Sciences Centre), project #2159; the Quebec Ministry of Health and Social Services multicenter ethics review (REB of Record: Comite d'ethique de la recherche du CHU Sainte-Justine), project #MP-21-2020-2863; the Weil Cornell Medicine General Institutional Review Board, protocol number 20-04021981; as well as the Comissao Nacional de Etica em Pesquisa, approval 4.305.792.

